# Tertiary Lymphoid Structure and CD8 T Cell Exclusion in Minimally Invasive Adenocarcinoma

**DOI:** 10.1101/2020.08.03.20166991

**Authors:** Jin Wang, Dongbo Jiang, Xiaoqi Zheng, Wang Li, Tian Zhao, Di Wang, Huansha Yu, Dongqing Sun, Ziyi Li, Jian Zhang, Zhe Zhang, Likun Hou, Gening Jiang, Fan Zhang, Kun Yang, Peng Zhang

**Affiliations:** Clinical Translational Research Center, Shanghai Pulmonary Hospital, School of Life Sciences and Technology, Tongji University, Shanghai, China; Department of Immunology, School of Basic Medicine, Air-Force Medical University. Xi’an, China; Department of Mathematics, Shanghai Normal University, Shanghai, China; Department of Thoracic Surgery, Shanghai Pulmonary Hospital, Tongji University School of Life Sciences and Technology, Shanghai, China; Tissue Bank, Department of Pathology, Experimental Animal Center, Shanghai Pulmonary Hospital, Tongji University School of Medicine, Shanghai, China; Academy of Medical Engineering and Translational Medicine, Tianjin University, China; Department of Gynecologic Oncology, Chinese PLA General Hospital, Beijing, China; Department of Thoracic Surgery, Shanghai Pulmonary Hospital, Tongji University School of Medicine, Shanghai, China

## Abstract

Lung adenocarcinoma is the leading cause of cancer death. To characterize the tumor microenvironment (TME) of early-stage lung adenocarcinoma, we performed RNA-seq profiling on 59 pairs of minimally invasive adenocarcinoma (MIA) tumors and matched adjacent normal lung tissues from Chinese patients. We observed mucin over-expression and glycosylation, and altered cytokine-cytokine interactions in MIA tumors, which also had distinct adaptive immune TME of higher CD4+ T cell infiltration, higher plasma B cell activation, and lower CD8+ T cell infiltration. The high expression of markers for B cells, activated CD4 T cells, and follicular helper T (Tfh) cells in MIA implicated the formation of tertiary lymphoid structures (TLS), which were supported by two independent single-cell RNA-seq data. Multiplex immunohistochemistry (mIHC) staining of 22 MIA tumors validated TLS formation and revealed an enrichment of follicular regulatory T cells (Tfr) in TLS follicles, which may explain the lower CD8+ T cell infiltration and attenuated anti-tumor immunity in MIA.

**Significance:** We discovered a potential immunosuppressive phenotype in MIA mediated by follicular regulatory T cells (Tfr) in TLS formation, which could be a potential mechanism of CD8+ T cell exclusion. Our study demonstrated how integrating tumor transcriptome and pathology can characterize the TME and elucidate potential mechanisms of tumor immune evasion.

## Introduction

Lung cancer is the world’s second-most common cancer but is by far the leading cause of cancer death. While most early lung cancer cases can be successfully treated, many lung cancers are not diagnosed until symptoms appear, at which time the disease has progressed to a more advanced stage. In recent years, low-dose computed tomography (LDCT) scan has been used for early lung cancer detection on people with a higher risk of lung cancer. Although LDCT has been shown to decrease the rate of lung cancer death, it also has a ∼20% false-positive detection rate and exposes people to low-dose radiation (1). While these imaging approaches are promising, early detection and treatment of lung cancer might be complemented by molecular approaches that characterize pre-cancerous lesions and their surrounding microenvironment. To better characterize the molecular, cellular, and tissue changes that drive tumor development and progression in its earliest stages, National Cancer Institute of the United States has recently issued new initiatives to construct an atlas of pre-cancer lesions and their surrounding microenvironment. Following The Cancer Genome Atlas (TCGA) project which successfully profiled thousands of human cancers, this new effort promises to provide better understanding of cancer initiation and bring new strategies for cancer diagnosis and treatment.

Non-Small Cell Lung Cancer (NSCLC) accounts for about 80% of all lung cancer incidences, with a majority belonging to the adenocarcinoma subtype. Early lung adenocarcinoma development is believed to progress from adenocarcinoma in situ, to minimally invasive adenocarcinoma (MIA), then to fully invasive adenocarcinoma (2). Despite the difficulty of diagnosing early lung cancer lesions and obtaining samples, there have been a number of genomics studies on early lung cancers in recent years (2-5). For example, one study examined the mutation patterns of in situ and invasive adenocarcinoma by whole-exome sequencing and compared the results with TCGA (2). Another study used targeted sequencing and droplet PCR to examine the mutation patterns in both early tumors and the matched blood samples (3). Most of these studies focused on identifying driver mutations potentially responsible for tumor initiation, which have been summarized in a recent review (6).

Besides tumor mutations, there has been growing appreciations of the important roles tumor microenvironment (TME) plays in cancer development, progression, and outcome. The TME consists of cancer cells, tumor-infiltrating lymphocytes, stromal and vascular cells, along with extra-cellular molecules such as cytokines and chemokines that modulate their interactions. Changes in cellular and extra-cellular components of the TME can facilitate tumor behaviors such as unchecked tumor growth and immune evasion (7). To examine such TME changes, gene expression profiles from RNA-seq are more informative than DNA-based tumor profiling.

Besides characterizing differentially expressed genes and pathways, RNA-seq has reasonable power to detect somatic mutations (8), gene fusions (9) and microsatellite instability (10), especially if matched normal samples are also available. There have also been multiple studies that use tumor RNA-seq data to infer immune cell populations and subtypes, and to associate immune infiltration with clinical outcomes or driver mutations (11,12). However, few such tumor profiling studies include matched adjacent tissues, and often the normal samples are sequenced at much lower coverage. One NSCLC study, which combined Whole Exome Sequencing (WES) and RNA-seq to discover the association between EGFR mutation and tumor-infiltrating B lymphocytes, did not profile RNA-seq from the matched normal, thus limiting the ability to characterize tumor-normal differences (13). Therefore, knowledge of the tumor immune microenvironment in early lung cancer patients, especially in comparison with the matched adjacent tissues, remains lacking.

In the TME, B cells and T cells are able to recognize and bind tumor antigens to induce adaptive immune responses (14,15). How cytotoxic T lymphocytes (CTL) influence immune response to tumor progression has been under intensive investigation in recent years (16). However, the role of B-T cell interactions in anti-tumor immune response has not been completely understood. Inflammation is known to be a potential cause and hallmark of early cancer development (17), and clusters of B and T lymphocytes can form tertiary lymphoid structure (TLS) at the site of inflammation(18). There has been increasing research interest in understanding TLS formation and function in human pathology, especially related to clinical implications and potential therapeutic targeting (19). A recent review summarized studies on the presence, composition, and functions of TLS in cancers and how TLS is correlated with therapeutic response (20). Three recent studies also reported TLS formation to be associated with response to immune checkpoint blockade (21-24). However, the impact of immune-stimulating or immune-suppressive components of TLS on tumor progression or immune evasion still awaits elucidation. Therefore, investigating TLS formation and its consequences on the tumor immune microenvironment in early stage tumors has the potential to inform disease etiology, precision medicine, and clinical prognosis.

In this study, we sought to characterize the TME of 59 pairs of minimally invasive adenocarcinoma (MIA) tumors and their matched adjacent normal tissues using RNA-seq. These tumors were initially diagnosed with LDCT, then surgically resected and confirmed for MIA by pathologists using hematoxylin and eosin (H&E) staining. To ensure an unbiased comparison between tumor and normal, we sequenced the RNA of both tumor and normal at higher depth than TCGA. This allowed us not only to examine the gene expression changes and infer immune cell infiltration levels, but also to compare the clonal expansion patterns of the immune receptor repertoires between the tumors and the matched normal tissues. Furthermore, by combining transcriptome data, H&E staining, and multiplex immunohistochemistry (mIHC) staining on paraffin sections, we confirmed the TLS formation in the MIA TME, which might be associated with immune suppression and tumor initiation. These data and results provide insights into the etiology of early lung adenocarcinoma, which may inform immunotherapy treatment strategies for lung cancers.

## Results

### Genes in cytokine-cytokine receptor interactions and mucin glycosylation are over expressed in MIA

We collected 59 pairs of pathology-confirmed MIA tumors and matched adjacent normal tissues **(Fig. S1, Table S1)**. All the MIA tumors were confirmed by pathology using the 2015 World Health Organization classification criteria of lung tumors (25). Hematoxylin and eosin (H&E) staining confirmed the tumor lepidic growth pattern and the less than 5 mm in diameter of invasive components **(Figure 1A)**. We conduced RNA-seq of all the samples at a minimum depth of 100 million paired-end 150bp fragments, roughly three times the sequencing depth of TCGA samples. We mapped the RNA-seq reads with STAR (26), quantified gene expression with HTSeq (27), and called differentially expressed genes between paired tumor and normal samples using edgeR (28). In total, we identified 1,471 up-regulated and 431 down-regulated genes with over 2-fold change and a false discovery rate (FDR) of less than 0.05 **(Table S2)**. We then applied gene set enrichment analysis (GSEA) (29) based on Kyoto Encyclopedia of Genes and Genomes (KEGG) (30) to examine altered pathways in MIA tumors **(Figure 1B)**. The most enriched up-regulated pathway in MIA is ABC-transporters which has an established tumor-promoting role in regulating cell survival, proliferation, migration, invasion, angiogenesis, and inflammation (31). In addition, widely studied cancer-associated pathways, such as Ras signaling and PI3K-AKT signaling (32), are also upregulated. Furthermore, MIA tumors exhibit stronger cytokine::cytokine-receptor interactions than adjacent tissues. One particularly overexpressed cytokine, *CXCL13*, encodes a chemokine that has been reported as a major B lymphocyte chemoattractant (33) **(Fig. S2)**. Another pathway showing significant enrichment in MIA tumors is mucin-type O-glycan biosynthesis. Many genes in the mucin family are over-expressed in MIA, especially *MUC21, MUC5B*, and *MUC4* **(Figure 1C)**, a phenomenon also observed in TCGA lung adenocarcinoma (LUAD) **(Fig. S3)**. Over-expression of mucin family proteins have been reported to contribute to the carbohydrate-enriched coating of cancer cells, which could promote integrin-mediated tumor growth and survival (34). In addition, overexpression and glycosylation of mucins, especially of MUC1, have been reported in multiple cancers and implicated to promote tumor metastasis and drug resistance (35,36). The overexpression of mucin family members likely accounts for the enrichment of mucin-type O-glycan pathway in MIA tumors.

**Figure 1.**
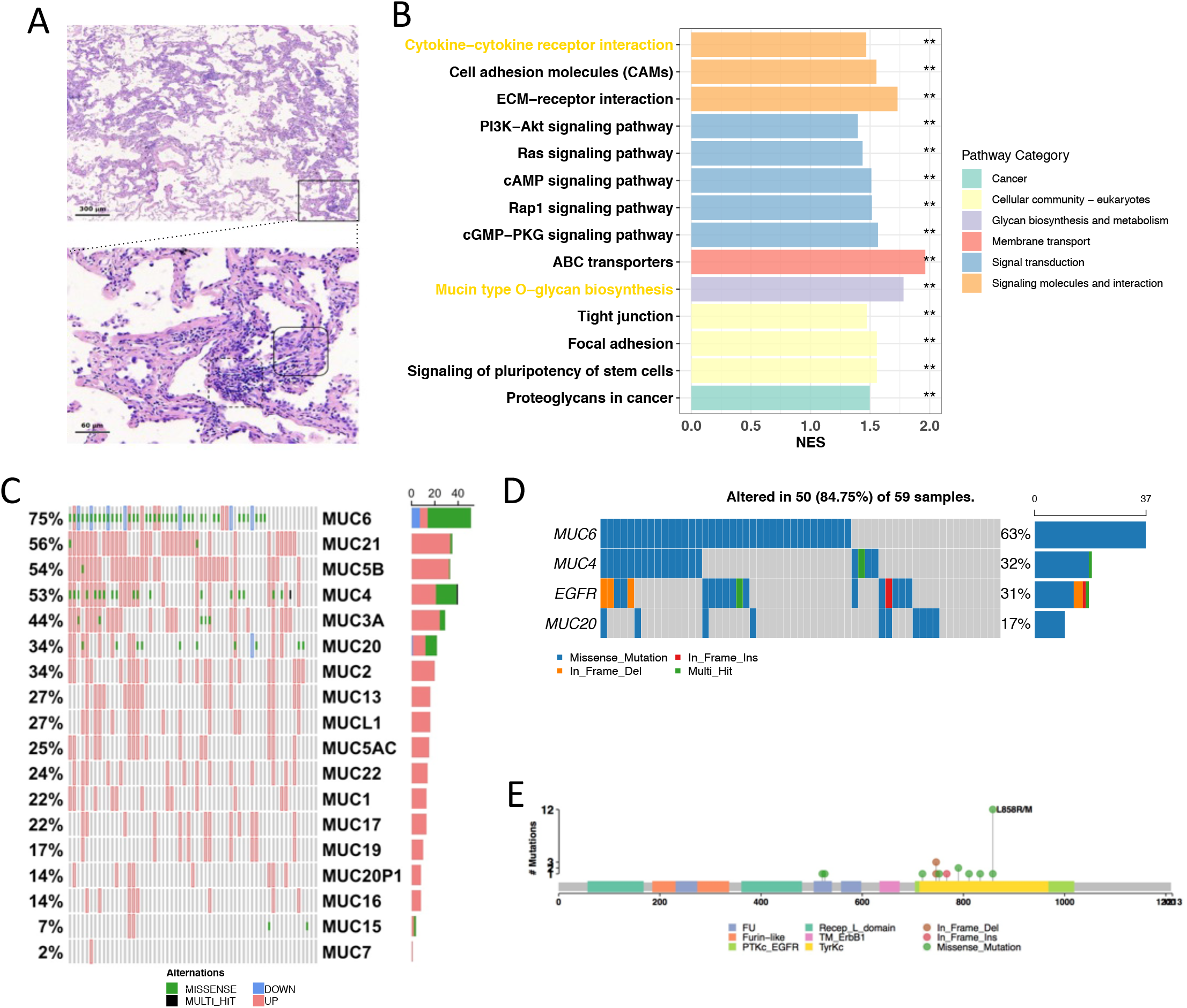
MIA tumors are characterized by highly mutated and over-expressed mucins, cytokine::cytokine-receptor interactions, and EGFR mutations. **(A)** Representative HE images of MIA. Top at 4X magnification, scale bar 300 µm. Boxed area of top image shown below (20x magnification, scale bar 50 µm). Lymphocyte infiltration (dashed box, bottom) and MIA tumor cells (solid box, bottom) are shown. **(B)** Normalized enrichment scores (x-axis) of tumor-upregulated KEGG pathways. Colors indicate different KEGG pathway categories. Asterisks denote significance level (*** FDR < 0.001; ** FDR < 0.01; * FDR < 0.05; NS FDR > 0.05). **(C)** Alterations of mucin genes in MIA patients. Each bar represents a MIA patient. Shown are mutation classifications, and expression status. Values on the right indicate the proportion of patients in which the gene was either mutated or had altered expression. **(D)** Most commonly mutated genes in MIA patients. Colors indicate variant type. Shown are the proportion of tumors containing a mutation in the indicated gene (left); total number of mutations in the indicated gene (right). **(E)** Schematic of the EGFR gene, showing mutations (stick and ball markers) and protein domains (color blocks).

### MIA tumors harbor EGFR and Mucin gene mutations

We next sought to detect mutations in MIA tumors by comparing RNA-seq data between tumor and adjacent normal tissue using Varscan2 (37). Although this is less sensitive than mutation calling from DNA-based approaches such as whole genome or whole exome sequencing, we still managed to find approximately 70 (median) variants per tumor, most of which were missense mutations **(Fig. S4A)**. The most enriched type of single nucleotide variants (SNV) is C>T transition **(Fig. S4B)**, which is known to be a signature of mismatch repair deficiency resulting in elevated microsatellite instability (MSI) (38). To computationally assess the MSI level of these MIA samples, we applied MSIsensor (10) to compare the RNA-seq data between tumor and normal. Although this is also less sensitive than DNA-based approaches, we indeed observed higher MSI score in MIA tumors with higher CT transition rate (**Fig. S4C**).

*EGFR* was found to be mutated in 31% of the MIA samples **(Figure 1D)**, consistent with the previous report of the prevalent *EGFR* mutations in progressive stages of lung adenocarcinoma (13). *EGFR* mutations occur mainly at the mutational hotspot “L858R/M” (39) **(Figure 1E)** in the kinase domain, in agreement observations in TCGA LUAD **(Fig. S4D)**. *EGFR* is known to participate in the signaling of the RTK-RAS pathway (32). The gain-of-function EGFR mutations primarily contributed to the oncogenic activation of the RTK-RAS pathway. Another important gene in the RTK-RAS pathway, *KRAS*, is mutated in only two tumors (**Fig. S4E**), and as expected, occurs mutually exclusively with *EGFR* mutations (40).

In addition to RTK-RAS pathway mutations, *MUC6* and *MUC4* genes were also highly mutated in MIA tumors. These two genes, as well as other MUC family members *MUC16, MUC5B* and *MUC17*, are frequently mutated in TCGA LUAD **(Fig. S3)**. Most of the MUC family SNVs are missense mutations, and together with their frequent over expression, suggest that tumor-associated mucins might create neoantigens in MIA tumors. We therefore applied OptiType (41) to infer the patient MHC Class I type and pVAC-Seq (42) to predict the likely neo-antigens in these MIA tumors. Indeed, our analysis supports mutated MUC6, MUC4, MUC20 as being potential neoantigens in MIA tumors (Fig. S5).

### Higher B cell and CD4+ T cell infiltration and activation but lower CD8+ T cell infiltration in MIA tumors

To decipher immune cell infiltration in the MIA TME, we estimated the immune cell composition from bulk tumor RNA-seq data using TIMER (43), CIBERSORT (11). Relative to adjacent normal tissues, MIA samples have significantly higher B cells and CD4+ T cells, and significantly lower CD8+ T cells and macrophages **(Figure 2A and Fig. S6)**. Consistent with the observation in MIA, TCGA LUAD tumors also showed higher B-cell and lower CD8+ T cell infiltration in tumors than the adjacent normal tissues. Interestingly, B cell infiltration gradually decreases towards normal during tumor progression from Stage I to Stage IV **(Figure 2B)** in TCGA LUAD, suggesting that increased B cell increase is associated with early LUAD onset.

**Figure 2.**
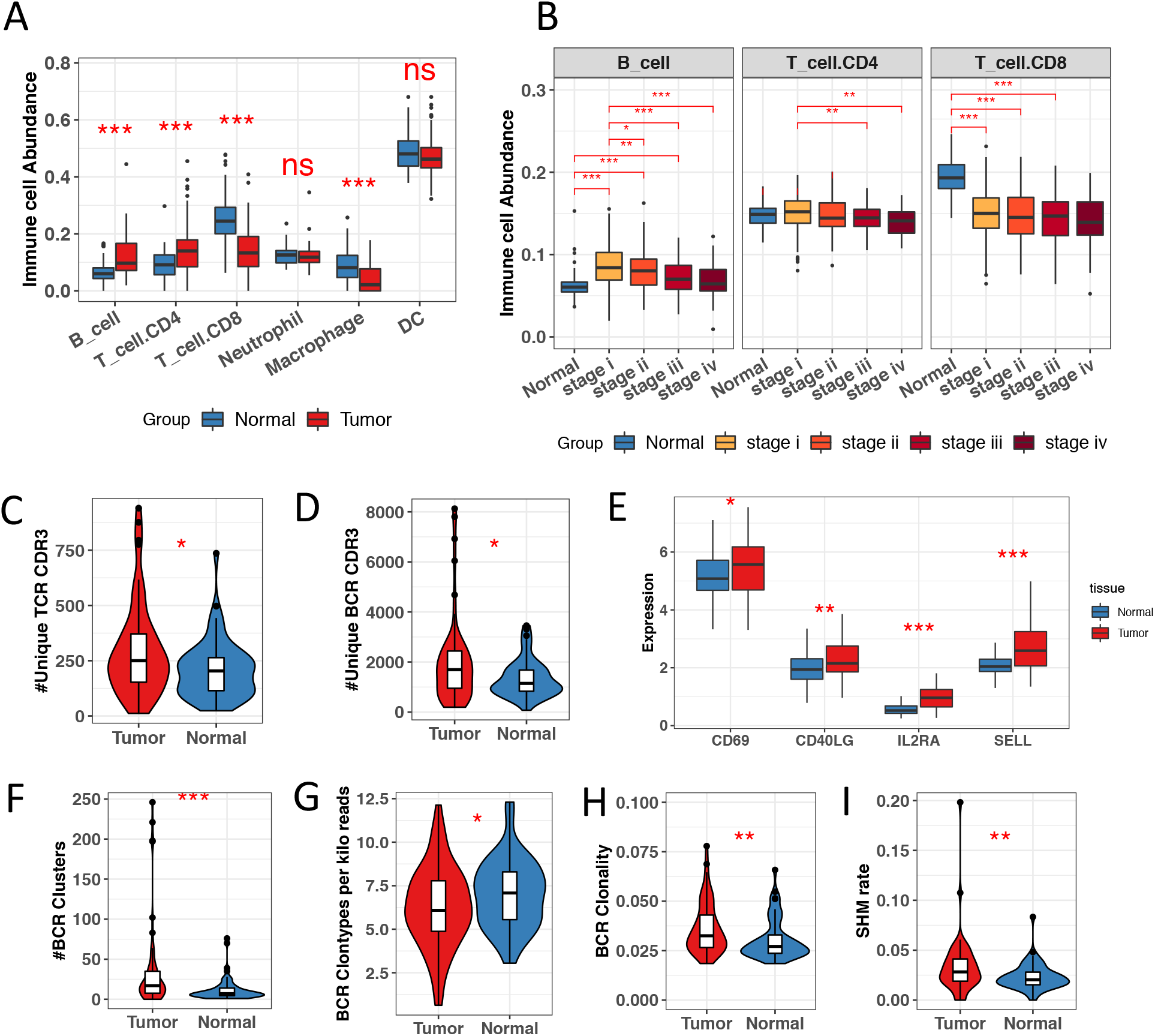
Higher B and CD4+ T cell infiltration and activation but lower CD8+ T cell infiltration characterize MIA tumors. **(A)** Comparison of immune components between tumor (red) and adjacent normal tissue (blue) in MIA. Immune Cell Abundance values (y-axis) were generated from TIMER. Significance levels are indicated (*** P < 0.001; ** P < 0.01; * P < 0.05; ns P> 0.05). **(B)** Immune infiltration among TCGA lung adenocarcinoma (LUAD) samples separated by clinical stage (colors). Comparison of fraction of TCR reads **(C)** and fraction of BCR reads (**D**) between MIA tumor and adjacent normal tissues. (**E**) Comparison of gene expressions on CD4+ T cell activation markers (CD69, CD40LG, IL2RA, and CD62) between MIA tumor and adjacent normal tissues. Comparison of the number of BCR clusters (**F**), BCR clonotypes per kilo reads **(G)**, BCR clonality **(H), and** somatic hyper-mutation rate of BCR **(I)** between MIA tumor and adjacent normal tissues. Asterisks denote significance on difference between MIA tumors and adjacent normal tissues (*** P < 0.001; ** P < 0.01; * P < 0.05).

Next we characterized the immune repertoires in these samples to investigate the activation and clonal expansion of tumor-infiltrating B and T cells. We ran TRUST, a computational algorithm we developed (44,45), to assemble the T-cell and B-cell receptor (TCR and BCR) repertoires from RNA-seq data. Significantly higher fractions of raw reads were mapped to TCR and BCR in MIA tumors than normal tissues (**Fig S7A-B**). Consistent with this observation, TRUST assembled significantly higher number TCR and BCR unique clonotypes of Complementary Determining Region 3 (CDR3) sequences in MIA tumors (**Figure 2C-D**). Since TRUST has better power to call the abundant and clonal-expanded TCRs and BCRs, the fact that TRUST assembled more TCR/BCR clonotypes already suggest that T cells and B cells have clonal expansion in the MIA tumors. Based on the CD4+ and CD8+ T cell infiltration trend in MIA vs adjacent normal, we expect most of the T cell activation and clonal expansion to occur in CD4+ T cells. Indeed, MIA tumors have significantly higher expression of CD4+ T cell activation markers CD69, CD40LG, CD25 (IL2RA), and CD62 (SELL) (46-49)(**Figure 2E**). However, since TCR diversity and clonality calculation could be confounded since we couldn’t distinguish TCRs from CD4+ vs CD8+ T cells, we turned to BCR to further evaluate immune cell clonal expansion. For B cells, we not only observed higher total number of BCR CDR3 clonotypes (**Figure 2D**) and BCR clusters (**Figure 2F**), but also lower BCR diversity (**Figure 2G**) and higher BCR clonality (**Figure 2H**) in MIA than adjacent normal, which strongly support B cell clonal expansion. In addition, B cells, upon antigen recognition and clonal expansion, undergo somatic hype-mutation (SHM) to evolve BCRs with better antigen binding affinity, and class-switch recombination (CSR) to transition to antibody-producing plasma cells. Indeed, we also observed higher BCR SHM rate (**Figure 2I**), higher CSR (**Fig S7C**) in MIA than adjacent normal, further supporting higher activated and clonal expanded B cells.

To check whether the activated tumor-infiltrating B and T cells could generate more diverse clonotypes for binding to different antigens in the TME, we compared the clonotypes detected between tumors and adjacent normal tissues. Majorities of the BCR (58.6%, **Figure 3A**) or TCR (57.8%) clonotypes were uniquely detected in tumors (**Figure 3B**), suggesting more B cell and T cell infiltration and clonal expansion in MIA tumors. We measured the repertoire similarity between each pair of MIA and adjacent tissue using Jaccard index (50,51), and found within-individual repertoire similarity to be much higher than the across-patient repertoire similarity (**Figure 3C**). Interestingly, the TCR and BCR similarities between tumor and normal (**Figure 3C**) are correlated, suggesting that T cells and B cells have coordinated clonal expansion in the MIA tumors. In addition, repertoire similarities among normal samples (N-N) are higher than that among MIA samples (T-T) from differential patients (**Figure 3D**), suggesting that tumor-specific antigens stimulate the expansion of different TCR and BCR clones in different tumors. Furthermore, tumors with higher mutation burden, as inferred from Varscan2, have significantly lower BCR (p-value < 0.01) and TCR (p-value < 0.001) repertoire similarity between tumor and adjacent tissues (**Fig. 3E**). Collectively, these observations suggest that TRUST-inferred immune receptor repertoires represent activated and clonal expanded B cells and potentially CD4+ T cells for recognizing and binding to tumor antigens in MIA.

**Figure 3.**
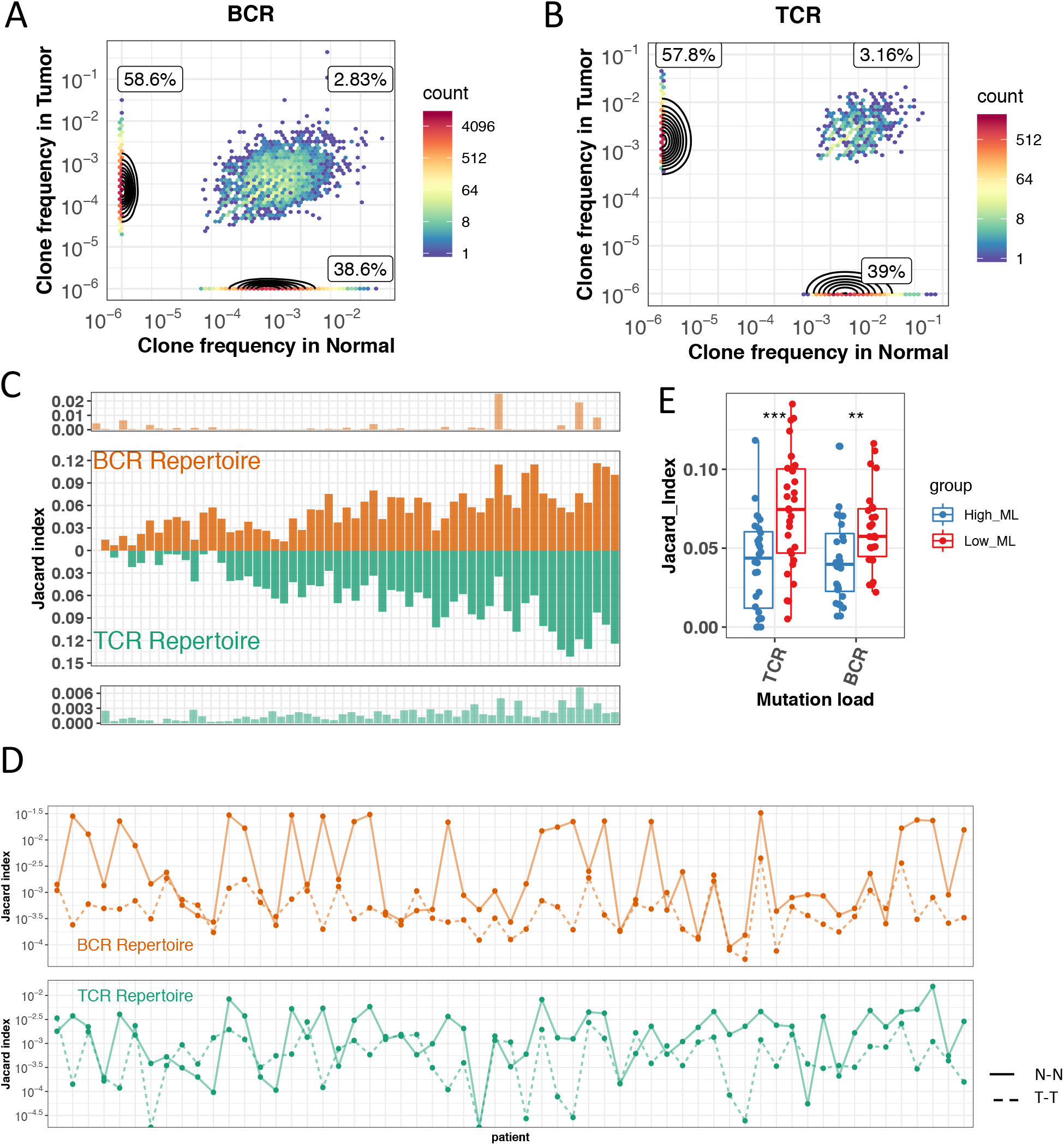
Tumor-infiltrating B cells and T cells are activated by tumor antigens in MIA. Normalized BCR clonotype frequency on patient level. Each dot represents a BCR clonotype and the normalized clonotype abundance in tumor and adjacent normal tissue. The three percentage values correspond to the overall percentage of tumor-specific clonotypes (58.6% in BCR), normal-specific clonotypes (38.6% in BCR), and shared (2.86% in BCR) clonotypes. **(B)** Normalized TCR clonotype frequency on patient level. **(C)** Jaccard similarity of the BCR/TCR clonotypes between tumor and adjacent normal tissues. Shown are shared clonotypes within each patient (dark brown for BCR, dark green for TCR) and average shared clonotypes between tumors and unmatched normal samples (light brown for BCR, light green for TCR). **(D)** Jaccard similarity of BCR and TCR repertoire among tumors (solid line) and normal tissues (dashed line) shown in top (BCR) and bottom (TCR) panel, respectively. **(E)** Jaccard similarity in groups with high (blue) and low (red) mutation load (*** P < 0.001; ** P < 0.01; * P < 0.05).

### Mucin mutation and overexpression is associated with CD4+ T cell-mediated B cell activation in MIA tumors

Since B cells could be activated by direct antigen stimulation and interaction with CD4+ T cells, we further checked how B cell activation was induced in MIA. It has been reported that MUC1 glycosylated peptides can induce MHCII-restricted B-T cell interactions (52). In addition, a synthetic peptide from the MUC1 tandem repeat region was shown to be presented by MHCII molecules which resulted in the initiation of T helper cell responses (53). In our MIA samples, we observed a positive association between mucin level and MHCII level (**Figure 4A**), raising the possibility that higher mucin expression in cancer cells might be associated with higher MHCII presentation. In addition, CD40 and its ligand CD40L are well-known to mediate the interaction between B cells and CD4+ T cells (54). We observed significant positive correlation between CD40 and CD40L in MIA tumors **(Figure 4B)**, which suggests CD4+ T cell-mediated B cell activation. The average level of CD40 and CD40L is associated with mucin expression (**Figure 4C**). Furthermore, we observed MIA tumors with over-expressed mucins have higher levels of IgG and IgA isotypes, suggesting higher level of B cell activation and transition to plasma cells (**Figure 4D**). Taken together, these results implicate the potential interactions between the mutation or overexpression on mucins, MHCII presentation to CD4+ T cells, and B cell activation and plasma cell transition in MIA tumors.

**Figure 4.**
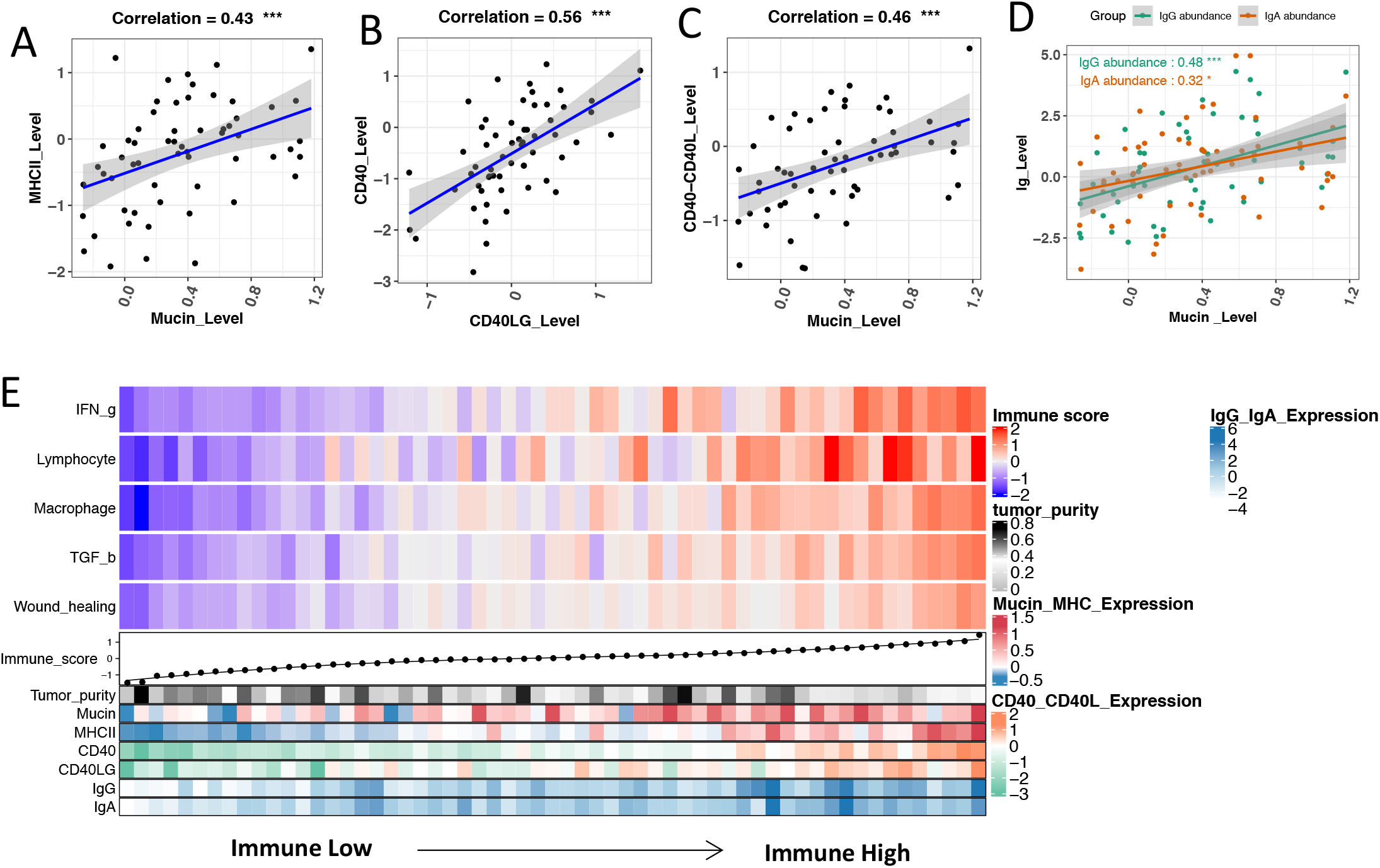
Mucin mutation and overexpression is associated with CD4+ T cell-mediated B cell activation in MIA tumors. The correlation between mucins and MHCII expression. **(B)** The correlation between CD40 and CD40LG expression. **(C)** The correlation between mucins and averaged level of CD40 and CD40LG. **(D)** The correlation between mucin expression with IgG (green) and IgA (brown) expression in MIA patients. **(E)** MIA tumors (x-axis) ranked by immune-related signature score (heatmap colors). The top bar shows the average immune signature score for each patient. The bottom bars show the tumor purity, expression level of mucins, MHCII molecules, CD40, CD40LG, IgG, and IgA.

To further investigate B cell activation among heterogeneous tumors, we characterized the overall immune status among the MIA samples using five signatures derived from the TCGA pan-cancer study (12). These signatures include “IFN-g” (representing IFN-g response), “Lymphocyte” (representing overall lymphocyte infiltration by B and T cells), “Macrophage” (representing the activation of macrophages/monocytes), “TGF-β” (representing TGF-β response), and “Wound healing” (representing core serum response). These five signatures seem to have correlated signals in the MIA tumors, so we used the average of the five to rank the MIA tumors from “immune-low” to “immune-high” **(Figure 4E, Methods)**. The “immune-high” tumors are associated with increased expression of MHCII (**Fig. S8A**) and mucins (**Fig. S8B**). In addition, the “immune-high” tumors are also associated with increased expression of CD40 (**Fig. S8C**) and CD40L (**Fig. S8D**), suggesting stronger interactions between B cells and CD4+ T cells. Furthermore, “immune-high” tumors are associated with higher B cell signals, such as IgG and IgA expression (**Fig. S8E-F**) and unique BCR clonotypes (**Fig. S8H**). Collectively, our observations suggest that mucin expression is associated with CD4+ T cell-mediated B cell activation in MIA adaptive immunity.

### Formation of tertiary lymphoid structure (TLS) in MIA tumors

Previous literature (18) reported that B-cell and CD4+ T-cell aggregation could lead to the formation of tertiary lymphoid structures (TLS). TLS starts from scattered B and T cell aggregation, progresses to follicles with separate T-Zones and B-Zones, and finally matures into ectopic germinal centers (EGC) with follicular dendritic cells (FDC) in the light-Zone for better B cell activation (20) (**Figure 5A**). In the EGC of mature TLS, B cells activates somatic hypermutations, isotype class-switches, and antibody production(19). So far our analysis revealed higher B-cell and CD4+ T-cell infiltration, lower CD8+ T-cell infiltration **(Figure 2A)**, and higher CD4+ T cell-mediated B cell activation **(Figure 3, Figure 4**) in MIA tumors. This motivated us to examine the expression of typical TLS markers for evidence of TLS formation in MIA. CD19 and MS4A1 (CD20), which are typical B-cell markers (55,56), were highly expressed in MIA tumors. The ligand-receptor pair CXCL13 and CXCR5, which are conventional indicators of follicles (57), were also significantly higher expressed in MIA tumors. Furthermore, the ligand-receptor pair CCL19 and CCR7, which are important for promoting T cell and B cell homing to lymphoid structures (58), were also highly expressed in tumors **(Figure 5B)**. These six TLS markers were all positively correlated with TIMER-estimated B cell and CD4+ T cell abundance but negatively correlated with CD8+ T cell abundance **(Figure 5B)**.

**Figure 5.**
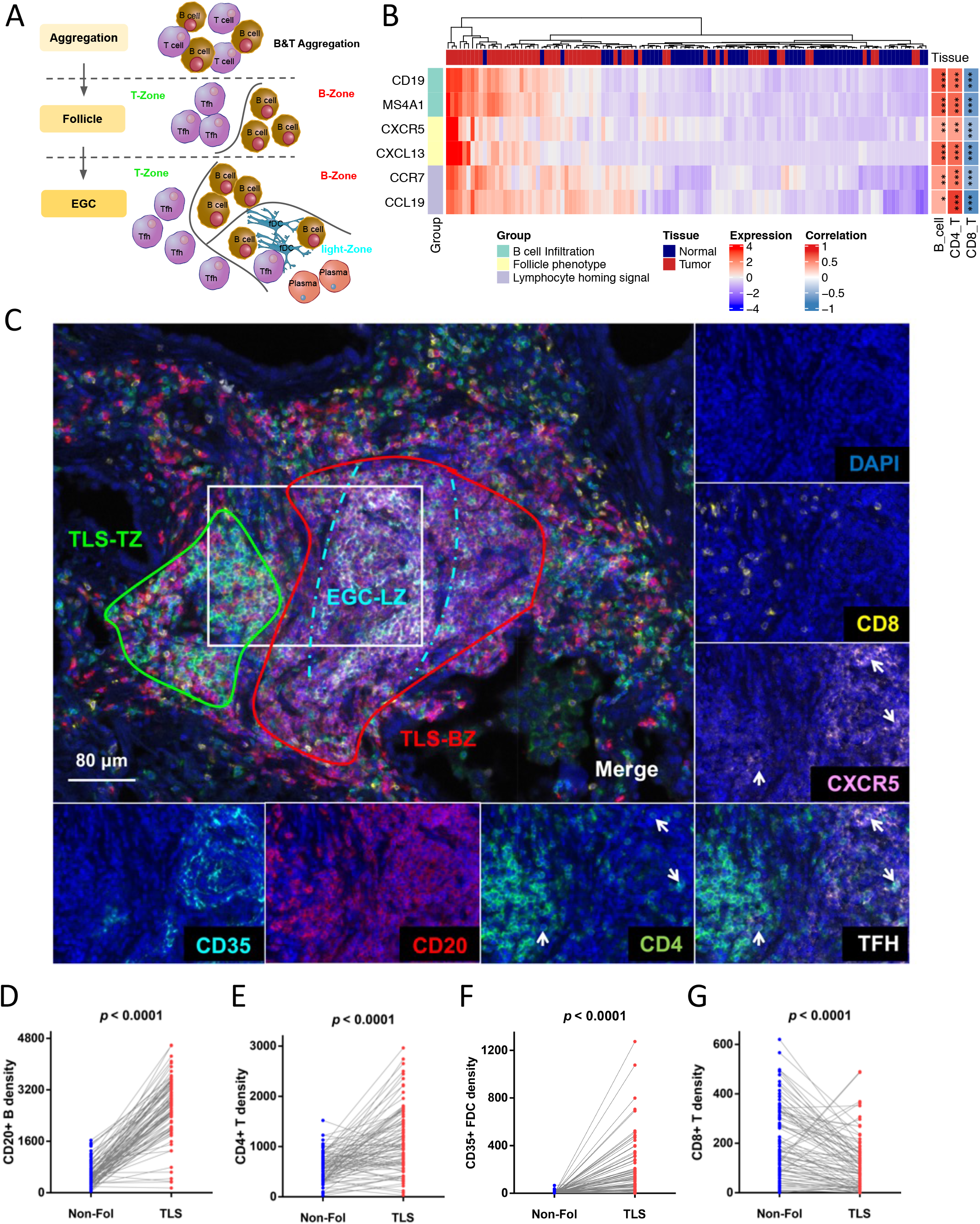
Formation of tertiary lymphoid structure (TLS) in MIA tumors. **(A)** The schema of TLS formation. The first stage is B & T cell aggregation. The second stage is follicle formation with T-Zone and B-Zone. The third stage is ectopic germinal center formation with formation with follicular dendritic cell in light-Zone. **(B)** Hierarchical clustering of Z-score normalized gene expression of B-cell infiltration markers (CD19, CD20), follicle phenotype markers (CXCR5, CXCL13), and lymphocyte homing signal markers (CCR7 and CCL19). Color of the top bar represents tumor (red) or adjacent normal (blue) tissue. Correlations between expression levels from these six TLS markers and immune cell abundance corrected by tumor purity are shown in the right bar. **(C)** Typical observation field of MIA six-plex TLS panel staining. The merged-channel multispectral image was captured after 5-cycle TSA staining and DAPI counterstain, revealing a classic lymphoid structure residing in MIA stromal tissue. Single channels are shown for the area bounded by the white rectangle. Fake colors are assigned as DAPI: blue, CD8: yellow, CXCR5: pink, CD4: green, CD20: red, CD35: cyan. Tfh cells are highlighted with white arrows (lower right corner images). Paired comparisons of CD20+ B cell density, CD4+ T cell density **(E)**, CD35+ follicle dendritic cell density **(F)**, and CD8+ T cell density **(G)** between Non-Follicle (Non-Fol) and TLS areas.

Besides the six TLS markers we evaluated above, another 12-chemokine gene signature has also been proposed to predict the presence of TLS (59,60). We observed highly correlated expression signals between our six TLS markers and the 12-chemokine signatures **(Fig. S9A)**. As a result, the two approaches give concordant TLS predictions in MIA tumors, and found TLS signals to be associated with immune-high tumors with stronger CD4+ T cell mediated B cell activation **(Fig. S9B-C)**. To further confirm the presence of aggregated B cells and CD4+ T cells in non-small-cell lung cancer (NSCLC), we also examined two recently published NSCLC single-cell RNA-seq datasets with tumors and adjacent normal tissues (61,62). Indeed, NSCLC tumors have higher level of MS4A1+ B cells, CD4+CXCR5+ follicular helper T cells (Tfh) than adjacent normal tissues (**Fig. S10A-B, Fig. S11A-B**). In addition, we used a computational algorithm CellPhoneDB (63) infer cell–cell communication from combined expression of multi-subunit ligand–receptor complexes to confirm the interaction between CD40LG in CD4+ T cells with CD40 in B cells (**Fig. S11C**). Collectively, our MIA RNA-seq data and NSCLC scRNA-seq data support TLS formation in lung adenocarcinoma (**Figure 4**).

With the transcriptome evidences of TLS formation in MIA tumors, we next sought to confirm TLS formation from pathology imaging. Firstly, we examined the hematoxylin and Eosin staining (H&E) pathology slides and observed lymphocytes aggregation **(Fig. S12)**, which support TLS formation in MIA tumors. Then, we used a six-marker multiplex immunohistochemistry (mIHC) to stain for TLS in 22 MIA tumors with high Tfh phenotype, which included the markers CD20 for B cells, CD4 and CXCR5 for follicular helper T cells (Tfh), CD8 for CD8+ T cells, and CD35 for follicular dendritic cells (FDC) **(Figure 5C)**. mIHC confirmed the formation of TLS in MIA tumors, although different samples showed varying level of TLS maturation **(Fig. S13)**. In the samples with mature EGC, we observed TLS B-cell zone (TLS-BZ) as marked by CD20, TLS T-cell zone (TLS-TZ) as marked by CD4, and EGC light zone (EGC-LZ) as marked by CD20, CD4, CXCR5, and CD35 **(Figure 5C, Fig. S14, Fig. S15)**. Furthermore, we segmented each observation field into different areas (Adenocarcinoma tissue, Non-Follicle, TLS T zone, TLS B zone, EGC light zone) to quantify and compare the immune cell density between TLS follicles and non-follicle areas. Here TLS follicle area includes TLS-TZ, TLS-BZ, and EGC-LZ, and non-follicle area includes scattered T cells and B cells outside of TLS. In MIA tumors, CD20+ B cells (**Figure 5D**, p-value < 0.0001), CD4+ T cells (**Figure 5E**, p-value < 0.0001) and CD35+ FDC (**Figure 5F**, p-value < 0.0001) were significantly higher in TLS than in non-follicular area. In contrast, CD8+ T cell showed significantly lower density (p-value < 0.0001) in TLS follicles than in non-follicle areas **(Figure 5G)**.

### Treg recruitment as potential mechanism of CD8+ T cell exclusion in tertiary lymphoid structures

Our transcriptome and mIHC data indicated that MIA tumors had lower CD8+ T cell infiltration (**Figure 2A**) and lower CD8+ T cell density in TLS (**Figure 5G**), respectively. In addition, MIA tumors have lower PD-L1 (CD274) expression than adjacent normal, suggesting PD-L1 independent mechanisms for CD8+ T cell suppression **(Fig. S16)**. This prompted us to explore the potential mechanism of CD8+ T cell exclusion in TLS and MIA. We examined immune suppressive cells, such as Tregs, M2 macrophages, neutrophils, monocytes, and fibroblasts (64) for possible cause of CD8+ T cell exclusion. Most suppressive cells were lower in MIA tumors than adjacent normal, with the exception of Treg cells (**Figure 6A**), suggesting a Treg-mediated CD8 T cell exclusion. From two independent scRNA-seq datasets, we also observed CD4+CXCR5+FOXP3+ follicular T regulatory (Tfr) cells to be enriched in NSCLC tumors (**Fig. S10B, Fig. S11B**). These data suggest that Treg cells might be responsible to suppress CD8+

**Figure 6.**
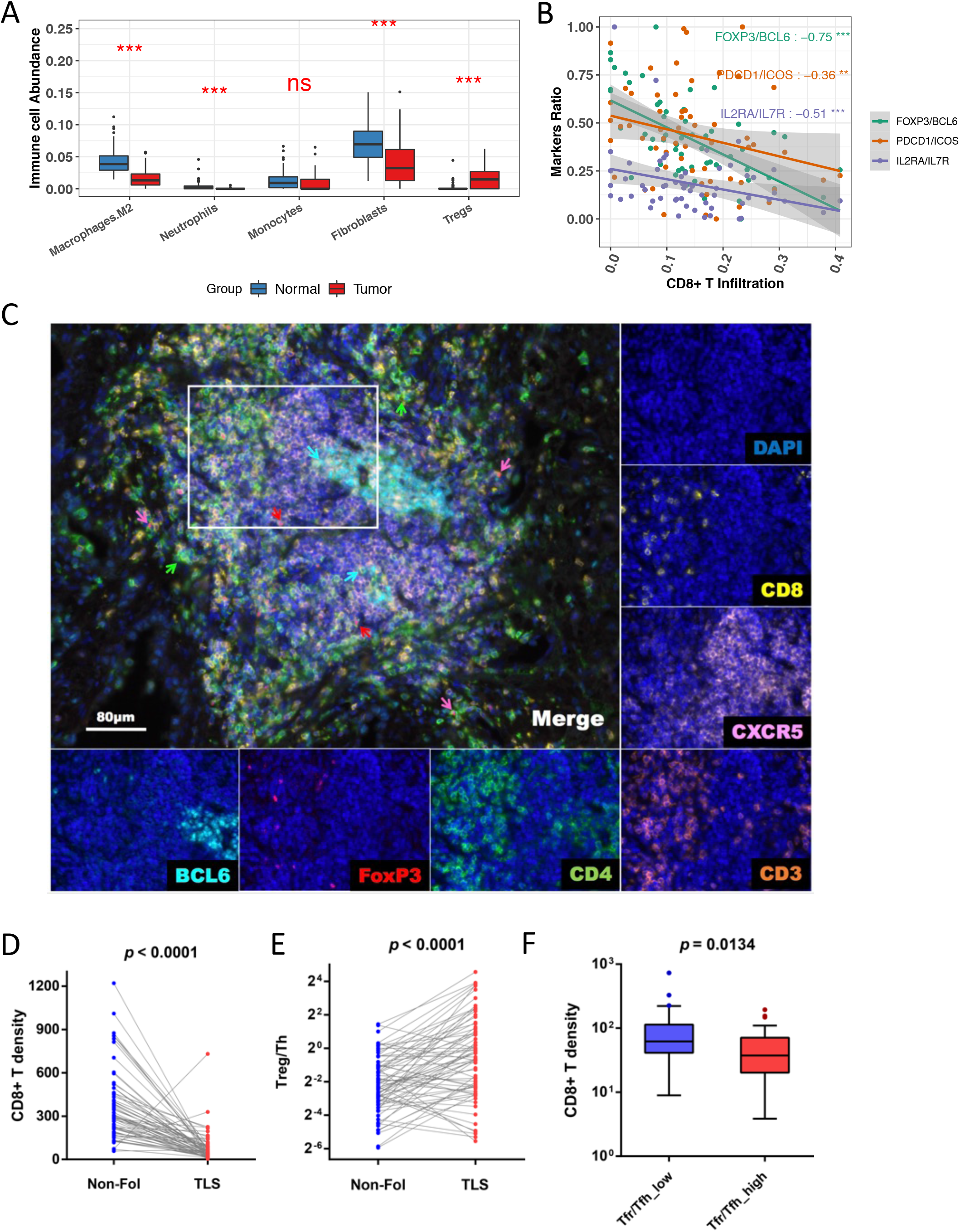
Treg recruitment as potential mechanism of CD8+ T cell exclusion in tertiary lymphoid structures. Comparison of immune suppressive cell infiltration estimated from XCELL between MIA tumor and adjacent normal tissues (*** P < 0.001; ** P < 0.01; * P < 0.05). **(B)** Plot of Tfr/Tfh marker ratio vs CD8+ T cell infiltration. FoxP3, PDCD1, and IL2RA are markers of Tfr cells, while BCL6, ICOS, IL7R are markers of Tfh cells. See text. **(C)** Typical observation field of MIA 7-plex panel staining for Tfr and Tfh. Merged multispectral image was captured after 6-cycle TSA staining and DAPI counterstain and reveals a mature TLS architecture. Single channels are shown for the area bounded by the white rectangle. Fake colors are assigned as DAPI: blue, CD8: yellow, CXCR5: pink, CD4: green, FOXP3: red, BCL6: cyan and CD3: orange. Sub-populations of CD4+ T-cells are highlighted by arrows (Tfr: red, Treg: purple, Th: green and Tfh: cyan). **(D)** Paired comparison of CD8+ T cell density between Non-Follicle (Non-Fol) and TLS areas. **(E)** Paired comparison of ratio of regulatory T cells to helper T cells between TLS and Non-Follicle areas. **(F)** Comparison of CD8+ T cell density between groups with high and low Tfr/Tfh ratio.

T cell infiltration in the region of lymphocyte aggregations **(Figure 4)**. To quantify the degree of immune suppression over immune stimulation, we measured the ratio of follicular regulatory T cells (Tfr) to follicular helper T cells (Tfh) in TLS by the normalized expression of three pairs of phenotype markers for Tfr and Tfh cells (FOXP3 over BCL6, PDCD1 over ICOS, and IL2RA over IL7R) (65), respectively. For each pair of markers, the ratio of Tfr / Tfh was negatively associated with CD8+ T cell infiltration **(Figure 6B)**, suggesting that Tfr in TLS might have negative effects on CD8+ T cell infiltration in MIA.

To further quantify the Tfr and Tfh in TLS, we used seven-marker mIHC staining on sequential slides of 22 MIA samples with high TLS expression markers. Specifically, we used CD3, CD4, CXCR5, and FOXP3 to detect Tfr cells and CD3, CD4, CXCR5, and BCL-6 to detect Tfh cells **(Figure 6C)**, respectively. Within the 69 available observation fields from the 22 MIA tumors, we found lower CD8+ T cell density in TLS follicles than non-follicular areas **(Figure 6D**, P < 0.0001), which was consistent with our observations in the six-plex mIHC panel **(Figure 5E)**. In addition, we observed a higher ratio of regulatory T cells to helper T cells in TLS follicles than in non-follicular areas **(Figure 6E**, P = 0.0032), indicating stronger immune suppression mediated by Tfr cells in TLS. To determine how the balance between immune suppression from Tfr and immune stimulation from Tfh influence CD8+ T cell infiltration in TLS, we split all the observation fields into two groups based on the Tfr / Tfh ratio. While higher Tfr / Tfh ratio was associated with lower CD8+ T cell density in TLS follicles **(Figure 6F)**, no such difference was found in non-follicular areas **(Fig. S17)**. These results suggest TLS formation to be associated with CD8+ T cell exclusion through Tfr mediated immune suppression.

## Discussion

In this study, we characterized the tumor microenvironment (TME) of minimally invasive adenocarcinomas (MIA) by integrating transcriptome data (molecular profiling, mutation analysis, immune cell estimation, and immune repertoire inference) and pathology staining (hematoxylin and eosin staining and multiplex immune-histochemistry). We observed varying levels of TLS formation, from the initial T cells and B cell aggregates, to follicle formation, to the mature ectopic germinal centers (EGC), in the MIA TME. Furthermore, we propose follicular Treg (Tfr) cells in the TLS as a potential CD8+ T cell exclusion mechanism in MIA tumors.

Our analysis revealed mucins in MIA samples to be highly mutated, over-expressed, and highly glycosylated. Mucins synthesized by epithelial cells are well-known to undergo glycosylation alterations during neoplasia (66-68). Aberrant glycosylation metabolism is common in cancers and known to facilitate cell survival and proliferation (66). Mucin overexpression may promote tumorigenicity and metastasis (66), which was proposed as an independent predictor for poor prognosis in lung adenocarcinoma (34). Glycosylated tumor-associated mucin peptides have been reported be bound by MHCII molecules and presented by B cells, leading to the activation of CD4+ T cells which in turn activates B cells through CD40-CD40L interactions (52,69). Our study identified higher tumor-infiltrating B cells and CD4+ T cells in both MIA and TCGA lung adenocarcinoma. Moreover, MIA tumors exhibited strong signals of B cell activation, somatic hyper-mutation and clonal expansion. Our observation of the positive associations of mucins with MHCII, B cell activation, and CD40-CD40L interactions potentially suggested mucin overexpression as a potential cause of CD4+ T cell-mediated B cell activation in MIA tumors. In addition, this CD4+ T cell-mediated B cell activation seemed to be associated with “immune-high” tumors.

Our observation of tumor-infiltrating B and CD4+ T cells suggested lymphocyte aggregation and TLS formation in the MIA TME. We therefore examined additional transcriptome and pathology evidences of TLS formation in MIA tumors. From MIA transcriptome, we observed up-regulation of six key TLS indicators (19,70) of B cell infiltration (CD19, CD20), follicle phenotype (CXCR5, CXCL13), and lymphocyte homing (CCR7 and CCL19). The results were corroborated using another 12-chemokine TLS signature (59,60). From pathology, we not only observed lymphocyte aggregation in the H&E slides, but also confirmed the co-localization of B cells, Tfh and FDC in TLS using six-panel mIHC. EGC are known to activate B cells for somatic hypermutation, antibody affinity maturation, class switching, and antibody production (71). We observed higher density of CD20+ B cells, CD4+ T cells, and CD35+ FDC, but lower density of CD8+ T cells in TLS follicles than in non-follicular areas. This motivated us to investigate whether cells previously reported to suppress CD8 T cell infiltration, such as Tregs, M2 macrophage, neutrophils, monocytes, or fibroblasts (64), inhibit CD8+ T cell infiltration in MIA. Further examination of bulk RNA-seq on our MIA tumors implicated Treg as the culprit of CD8 T cell exclusion in MIA tumors. This hypothesis was supported by previous auto-immune studies in human and mouse (72-76). Using mIHC to evaluate the ratio between immune suppression (Tfr) and immune stimulation (Tfh), we found a negative association between the Tfr / Tfh ratio and CD8+ T cell infiltration, further supporting Tfr-mediated CD8+ T cell exclusion in MIA.

Controversy still exists regarding the impact of TLS on clinical prognosis. For example, TLS was reported to be associated with poor prognosis in human hepatocellular carcinoma (77) but good prognosis in colorectal cancer and breast cancer (78,79). In NSCLC cohorts, TLS in the tumors was observed to be associated with favorable prognosis (80-82), dependent on high CD8+ T cell and mature dendritic cell infiltration in the tumors, although we observed a negative association between TLS with CD8 T cell infiltration in MIA tumors. Intriguingly, recent studies also reported TLS to be associated with better response to immune checkpoint inhibitors in melanoma and sarcoma (21,22,24). These studies suggest that the discrepancies of TLS on clinical prognosis may be dependent on the cancer type and clinical contexts. Our observation that TLS formation could be associated with Tfr recruitment, CD8 T cell exclusion, and immune suppression have potentially important implications in cancer immunotherapy: while tumors with TLS and a predominant Tfh recruitment might directly respond to anti-PD1 treatment, those with TLS and a predominant Tfr recruitment might benefit from initial anti-CTLA4 treatment to deplete the Tregs before responding to anti-PD1 treatment. Our current study is still limited in sample size and statistical power, although another study on medRxiv reported similar findings in early lung cancers (83). Currently, our analysis only revealed associations without having functional validation of the causal effects. Therefore, future research and clinical studies are needed to fully elucidate the mechanism of CD8 T cell exclusion and explore better immunotherapy strategies in cancer.

## Methods

## Sample collection and RNA sequencing

All 118 samples were collected from 59 MIA patients at Pulmonary Hospital, Shanghai, China. All specimens were obtained from patients with appropriate consent from the relevant institutional review board. The clinical information of these patients was shown in **Table S**1. The RNA extraction procedure was compiled with standard protocols. DNA and RNA were collected from samples using AllPrep DNA/RNA Mini Kit (Qiagen, 80204, Germany). The cDNAs were synthesized and used to construct RNA libraries using NEBNext Ultra RNA Library Prep Kit (NEB, E7530, USA) and NEBNext Multiplex Oligos for Illumina (NEB, E7335, USA). The libraries were sequenced on the Illumina HiSeq2500 platform via a 2×150bp paired-end sequencing at Annoroad Gene Technology (Beijing, China). For each sample, more than 200M fragments were sequenced.

### Gene profile processing of MIA RNA-Seq data

The transcriptome data were processed by the standard RNA-Seq pipeline. We performed sequence mapping by STAR alignment (26), reads quantifying by HT-Seq Count (27), and gene expression measurement by TPM converting from read count. Differential expression analysis on paired samples was performed using edgeR (28). Gene set enrichment analysis was performed using GSEA (29). Pathway categories are referred from KEGG website(https://www.genome.jp/kegg/pathway.html)

### TCGA data downloading

For TCGA LUAD dataset, we downloaded gene expression data and mutation data from firebrowse (http://firebrowse.org).

### Mutation calling from paired RNA-Seq data

Mutation detection from paired RNA-Seq data was analyzed by Varscan2. Somatic variants from Varscan2 (37) were annotated by Variant Effect Predictor(VEP) (84) and vcf2maf (https://github.com/mskcc/vcf2maf). Variants with low quality were removed based on the criteria which GDC pipeline recommends (https://docs.gdc.cancer.gov/Data/File_Formats/MAF_Format/). Mutation visualization was implemented by Maftools (85) and Complexheatmap (86).

### Normalization for altered mucin expression

Z-score normalization of mRNA expression for patients was performed. One over-expressed or down-expressed gene was expected to have a difference of at least two standard deviations from the mean of expression in the reference samples.

### Immune infiltration estimation from RNA-seq data

We applied Tumor IMmune Estimation Resource (TIMER) (43) to deconvolve the immune cell components in the TME using TPM expression data. To ensure robust abundance estimation, CIBERSORT (11) according to the predefined LM22 signature and XCELL (87) were both used to estimate immune cell infiltration.

### The immune signature score for classifying “immune-low” and “immune-high” MIA tumors

From the pan-cancer paper (12), the immune signature data and the pre-trained scoring model were downloaded from the CRI-iAtlas (https://github.com/CRI-iAtlas/shiny-iatlas/). Based on the model, we computed the five representative immune signatures score from the expression data, which was normalized by paired normal tissues and median-centered by gene level. Then, we ranked MIA tumors from immune “low” to immune “high” based on the averaged immune signature score.

### Definitions of BCR repertoire metrics

We applied TRUST3 to infer the immune repertoire from bulk RNA-seq data (44,45).We measured basic metrics to quantify immune repertoire and further performed paired comparison of TCR and BCR repertoire between tumor and tumor-adjacent tissues. The fraction of TCR or BCR reads was defined as the library size located in BCR loci, which is normalized by total mapped reads (E1). The number of unique TCR or BCR CDR3 sequences summarized the total unique CDR3 size. The number of TCR or BCR clonotypes per kilo reads measures TCR or BCR diversity. In addition, B cells undergo clonal expansions with a high rate of somatic hypermutation by single base substitutions in CDR3 sequences. Thus, we built BCR lineages referred to the method of a previous study (45), which generates 8-mer CDR3 motifs and an upper triangular matrix of counting the substitutions between DNA sequences from any two clonotypes in one BCR lineage to cluster partial and complete CDR3 sequences. For each sample, BCR or TCR clonality is defined by Shanno entropy (E2) (88). Somatic hypermutation rate was defined as the total count of harboring one base substitution normalized by the total length of aligned DNA sequence (E3). Ig class switch recombination (CSR) were built for the BCR clonotypes with different Ig isotypes from the same BCR lineage. Any two Ig isotypes within the same cluster motif could be switched based on the specific order, which is IgM, IgD, IgG3, IgG1, IgA1, IgG2, IgG4, IgE, IgA2 (89,90). Here we only remain the Ig isotypes with abundance more than 1% and check CSR on IgG and IgA for measuring B cell activation signal. The ratio of CSR was computed referring to the method in the previous study (45). The equations are shown below.

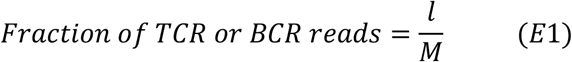

*l* denotes library size located in BCR loci; *M* denotes total mapped reads.

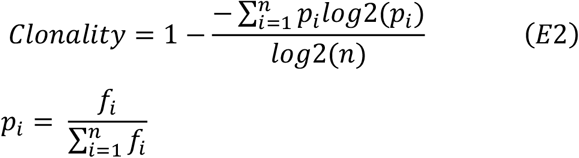

*n* denotes total number of TCR clonotypes or total number of BCR clusters and non-clustered clonotypes, *f*_*i*_ denotes clonal frequency for non-clustered clonotypes or clustered clonotypes.

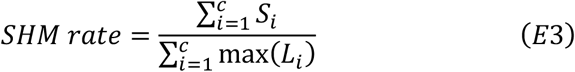

*c* denotes the number of BCR clusters, *S*_*i*_ denotes the sum of clonotype pairs harboring one base substitution, *L*_*i*_ denotes the max number of aligned non-gapped DNA sequence in one cluster.

### Tumor purity evaluation

Tumor purity was estimated by the algorithm of EPIC (91) (http://epic.gfellerlab.org), which is designed for estimating the absolute fraction of immune cells and cancer cells simultaneously from RNA-seq expression data. We selected “tumor infiltrating cells” as reference profile to predict cell fractions.

### Single-cell RNA-seq data analysis

Two independent single-cell RNA-seq datasets were collected from NSCLC cohorts with tumors and adjacent normal tissues (GSE139555, EMTAB6149). Single-cell RNA-seq dataset pre-processing was performed by the comprehensive pipeline MAESTRO (92). Single cell clustering and cell type annotation are available to be visualized in TISCH (http://tisch.comp-genomics.org/). In the dataset of NSCLC_ GSE139555, only cells from tumor and normal tissues were extracted from raw count data for comparing gene expression of cells between tumor and normal. In the dataset of NSCLC_ EMTAB6149, cluster averaged expression data was downloaded from the paper supplementary data due to no available tissue information on cell barcode level. Cell-cell interaction analysis was inferred utilizing a public method CellPhoneDB (63). Only receptor-ligand pairs significantly enriched in less than 5% of all cell-cell interactions were exhibited for the cell interactions on B cells or T cells in the dot plot.

### Immunohistochemistry

Slides at 4-μm thickness were cut from paraffin blocks from MIA biopsies (n=59) and paired normal lung tissue. Slides were then stained for the interested markers in certain condition of antigen retrieval and incubation (**Table S3**) with Dako Real kit according to IHC-P protocol. Tonsil tissue were performed as control for staining optimization.

### Multiplex immune-histochemistry staining for lymphocyte infiltrating in MIA

Twenty-two matched paraffin blocks of the 59 samples were processed through the 4-μm-thick section, deparaffinization in xylene, rehydration in an ethanol gradient. Slides were stained according to Opal 7-plex technology (PerkinElmer), yielding to the simultaneous visualization of seven markers (containing DAPI for nucleus) on the same slide. During each of the six cycles of staining, antigen retrieval (AR) was conducted via microwave treatment in AR solution pH 6 or pH 9 (AR6 or AR9) suggested by IHC validation; blocking was followed by instructions for 15 mins at room temperature (RT); and primary antibodies were then incubated for 1 hr at RT or overnight at 4 °C. Next, HRP labeled polymer goat anti-mouse and rabbit antibodies were incubated at RT for 10 min followed by TSA opal fluorophores (Opal 520, Opal 540, Opal 570, Opal 620, Opal 650 or Opal 690) 10 min incubation. Microwave treatment was performed to remove the antibody TSA complex at each cycle of staining with AR solution (pH 9 or pH 6). Finally, all slides were counterstained with DAPI for 5 min and enclosed in ProLong Antifade Mountant (Solarbio). The slides were scanned using the PerkinElmer Mantra System, and the multispectral images obtained were unmixed using spectral libraries that were previously built from images stained for each fluorophore (mono-plex), using the inForm Advanced Image Analysis software (inForm 2.4.1 PerkinElmer).

### Multispectral image analysis for tumor microenvironment of MIA

The entire area of the sections was stained, scanned, scored, and quantified. The scoring protocol consisted of several automated steps: tissue categorization, cell segmentation, and cell phenotyping. Using the integrated InForm image analysis software, multispectral images that were representative of different samples were selected and used to train the InForm software for categorization of the tissue into epithelium and stroma. Microinvasive epithelium without infiltration was tagged as “ACC” for cancer cell of adenocarcinoma, while lymphocyte infiltrating stroma area was further recognized as “Non-Fol”, “TLS-BZ” and “TLS-TZ”, representing scattered lymphocytes infiltration, B-cell zone of TLS formation, and T-cell zone of TLS formation. The settings learned from the training on the representative images from different samples were saved within an algorithm, which enabled a batch analysis of all the tissue slides. We designed six-plex TLS panel for TLS component cells and seven-plex Tfr/h panel for suppressive/stimulatory immune cells. They were used on two sequential slides to characterize the immune microenvironment of MIA lesions. The TLS panel included CD4, CD8, CD20, CD35, DAPI, and CXCR5 while the Tfr/h panel included CD3, CD4, CD8, FOXP3, BCL-6, DAPI, and CXCR5. The software classified the tumor (epithelial) and infiltrated stromal regions. The classification was verified and approved by a certified pathologist (W.D.). For six-plex mIHC panel, CD20+ B density is defined as the percentage of CD20+ cells in total immune cells, including CD8+ cells, CD20+ cells, CD35+ cells, and CD4+ cells. Similarly, CD4+ T density is defined as the percentage of CD4+ cells in total immune cells. CD35+ density is defined as the percentage of CD35+ cells in total immune cells and CD8+ T density is defined as the percentage of CD8+ cells in total immune cells. For seven-plex mIHC panel, we defined the regulatory T-cell phenotype with CD3^+^CD4^+^CXCR5^+^FOXP3^+^Tfr and CD3^+^CD4^+^CXCR5^−^FOXP3^+^Treg and stimulatory T-cell phenotype with CD3^+^CD4^+^CXCR5^+^FOXP3^−^Tfh and CD3^+^CD4^+^CXCR5^−^FOXP3^−^Th. Either cell densities or absolute counts were enrolled for calculation.

### Statistical analysis

For all the statistical test for comparing the difference between tumor and match adjacent tissues, Mann–Whitney U test was used to evaluate the P values and P< 0.05 was considered statistically significant. Multiple hypothesis correction was computed in the form of adjusted p-value through the Benjamini-Hochberg procedure. The thresholds used in the differential expression analysis were 2.0 for fold-change and 0.05 for the adjusted P value. Spearman’s rank correlation coefficient was presented as the association of two independent features. A partial correlation test was used to measure the correlation between genes and immune cell infiltration when corrected by tumor purity. All the statistical tests were implemented using the R programming language.

## Data Availability

The RNA-seq data is depositing in Genome Sequence Achive (GSA) of National Genomics Data center (NGDC). All the data will be available in the manuscript or from the authors upon request

## Authors’ Contributions

**Conception and design:** Jin Wang, Dongbo Jiang

**Development of methodology:** Jin Wang, Dongbo Jiang

**Acquisition of data (acquired and managed patients, provided facilities):** Peng Zhang, Gening Jiang, Fan Zhang, Wang Li, Tian Zhao

**Analysis and interpretation of data (e**.**g**., **statistical analysis, computational analysis, experiment analysis):** Jin Wang, Dongbo Jiang, Xiaoqi Zheng, Jian Zhang, Dongqing Sun, Zhe Zhang, Kun Yang

**Writing and review the manuscript:** Jin Wang, Dongbo Jiang, Ziyi Li, Kun Yang

**Administrative, technical, or material support:** Huansha Yu, Di Wang

**Study supervision:** Peng Zhang, Kun Yang

## Data Availability

The RNA-seq data is deposited at Genome Sequence Archive (GSA), which is hosted by National Genomics Data Center (NGDC) under the submission code: subCRA003749. All the other data could be found at manuscript or available from the authors upon request.

## Acknowledgments

The authors acknowledge Dr. X. Shirley Liu in Dana-Farber Cancer Institute and Dr. Arlene Sharpe in Harvard Medical School for the helpful discussion and advice, to Drs. Jia Liang and Rongjian Su in Jinzhou Medical University, Mr. Wei Liu and fellows of the SKLBCE-PerkinElmer Center of Excellence, Beijing, China for their assistance on the mIHC experiments.

This work was supported by the National Natural Science Foundation of China (Grant No. 81872290, 81972172, and 81772763), the Shanghai Science and Technology Committee (No. 19XD1423200, 18140903900).

## FIGURE LEGEND

**Figure S1.**
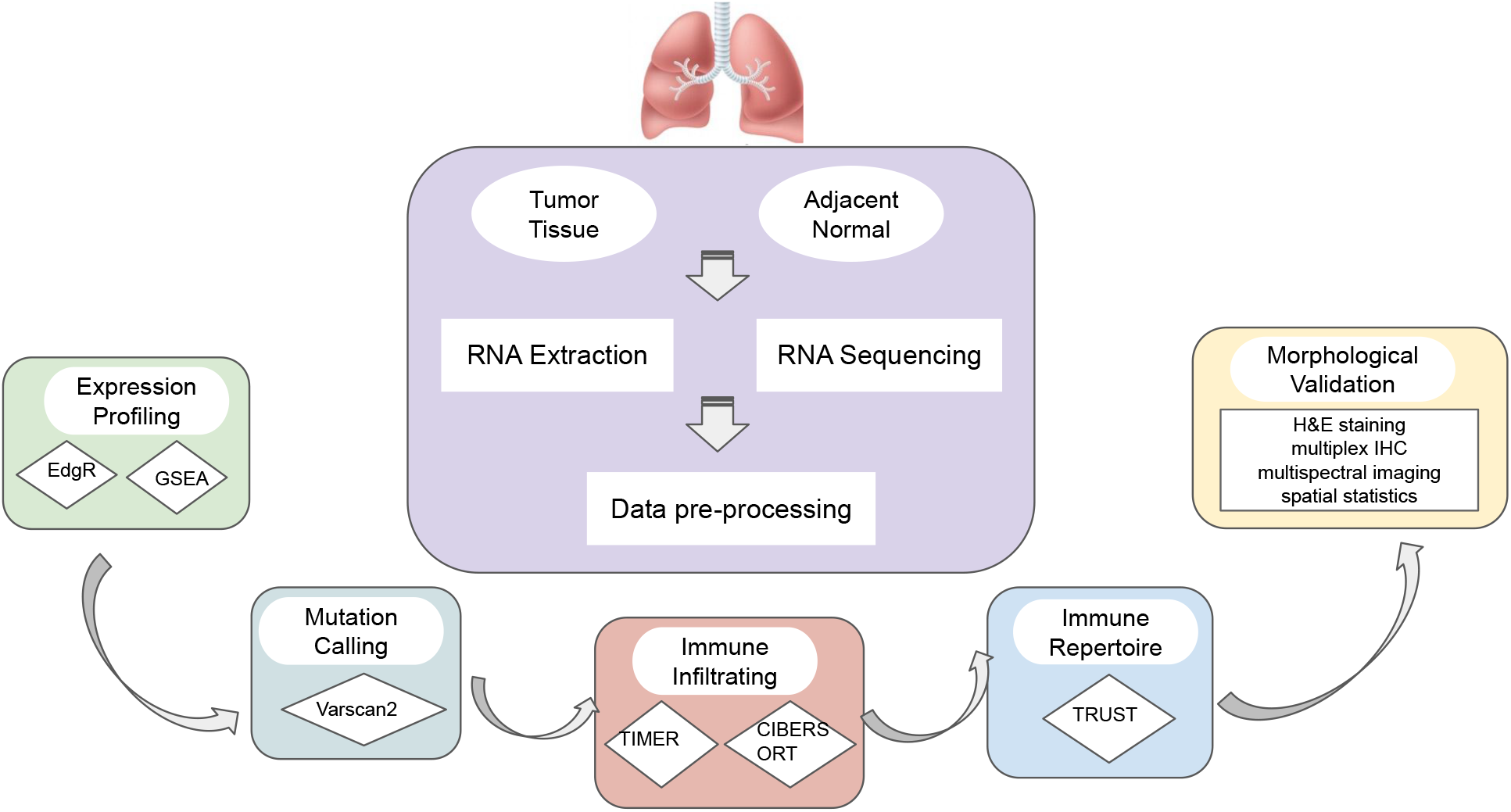
The workflow of data processing. In this workflow, five sections were performed for analyzing transcriptome data and imaging data, including expression profiling EdgR and GSEA, mutation calling from Varscan2, estimating immune cell infiltration from TIMER and CIBERSORT, quantifying immune repertoire from TRUST3, and Morphological validation from H&E staining, multiplex IHC, multispectral imaging and spatial statistics.

**Figure S2.**
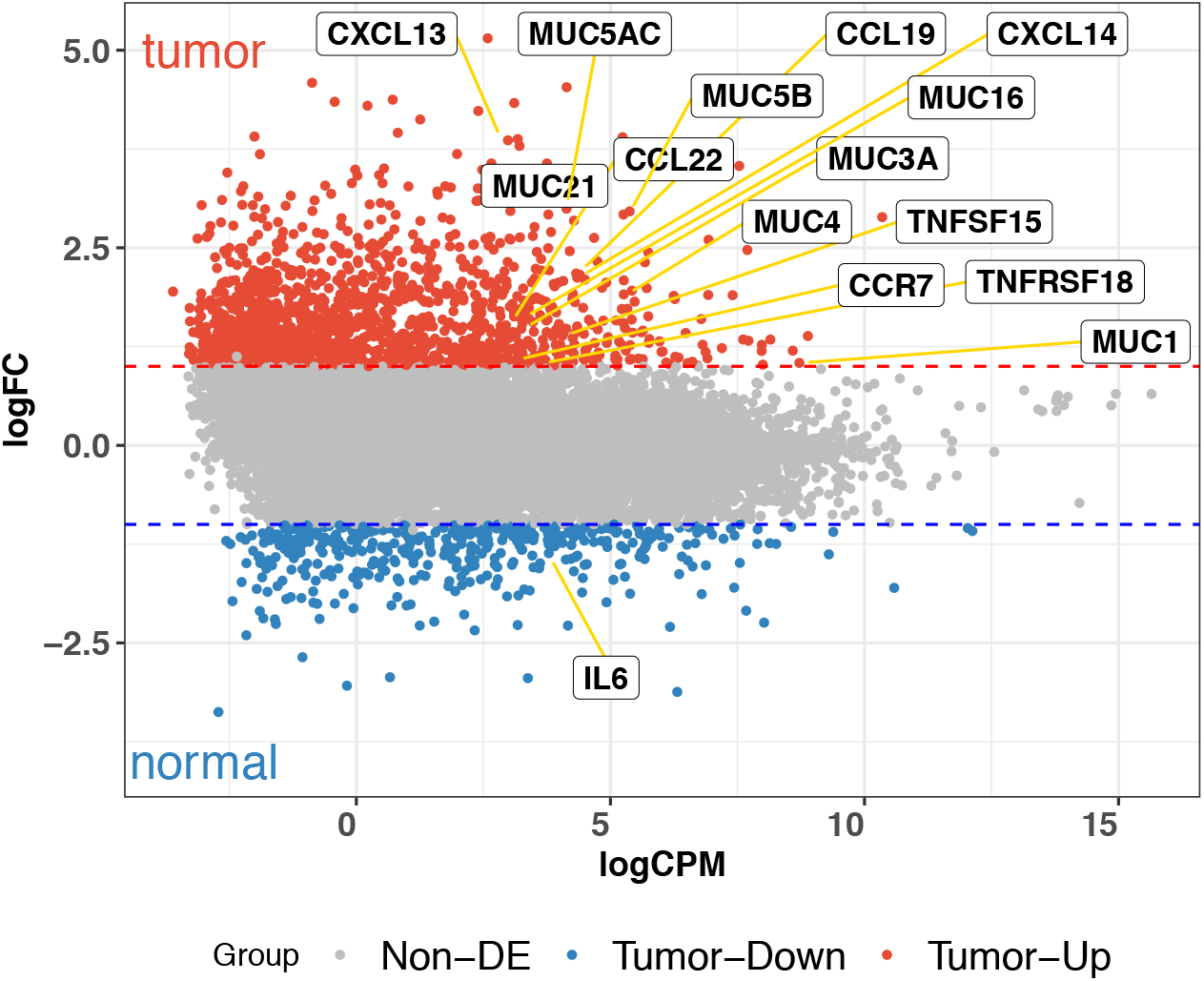
Differential gene expression analysis. Colors of dots represent the tumor up-regulated genes (red) and tumor down-regulated genes (blue). Top differentially expressed mucin family members, cytokines, and cytokine receptors are highlighted in Volcano plot.

**Figure S3.**
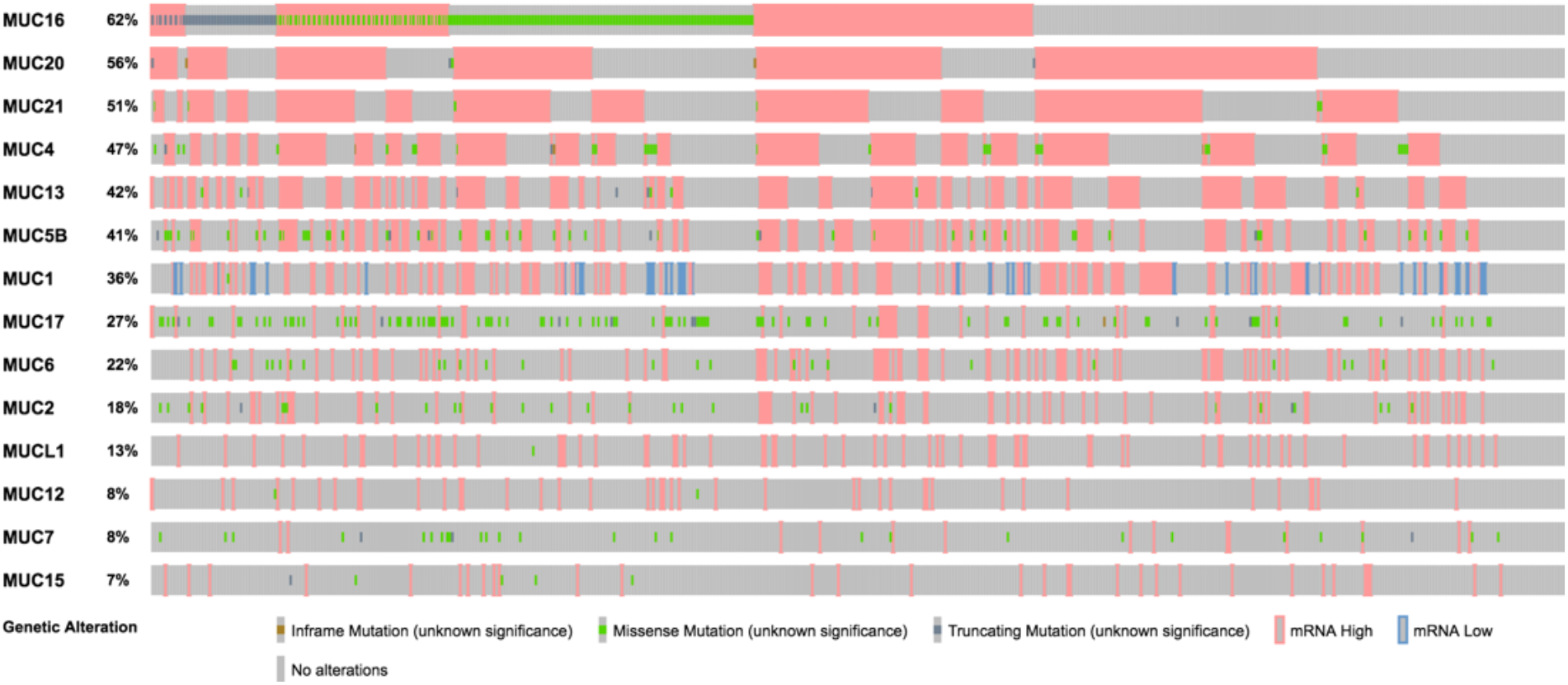
Genetic alterations of mucin family members in TCGA LUAD. Mutations and gene alterations in mucin family members are highlighted with different colors, including inframe mutation (pink inside), missense mutation (green inside), truncating mutation (blue inside), mRNA high expression (pink outside), mRNA low expression (blue outside), and no alterations (grey).

**Figure S4.**
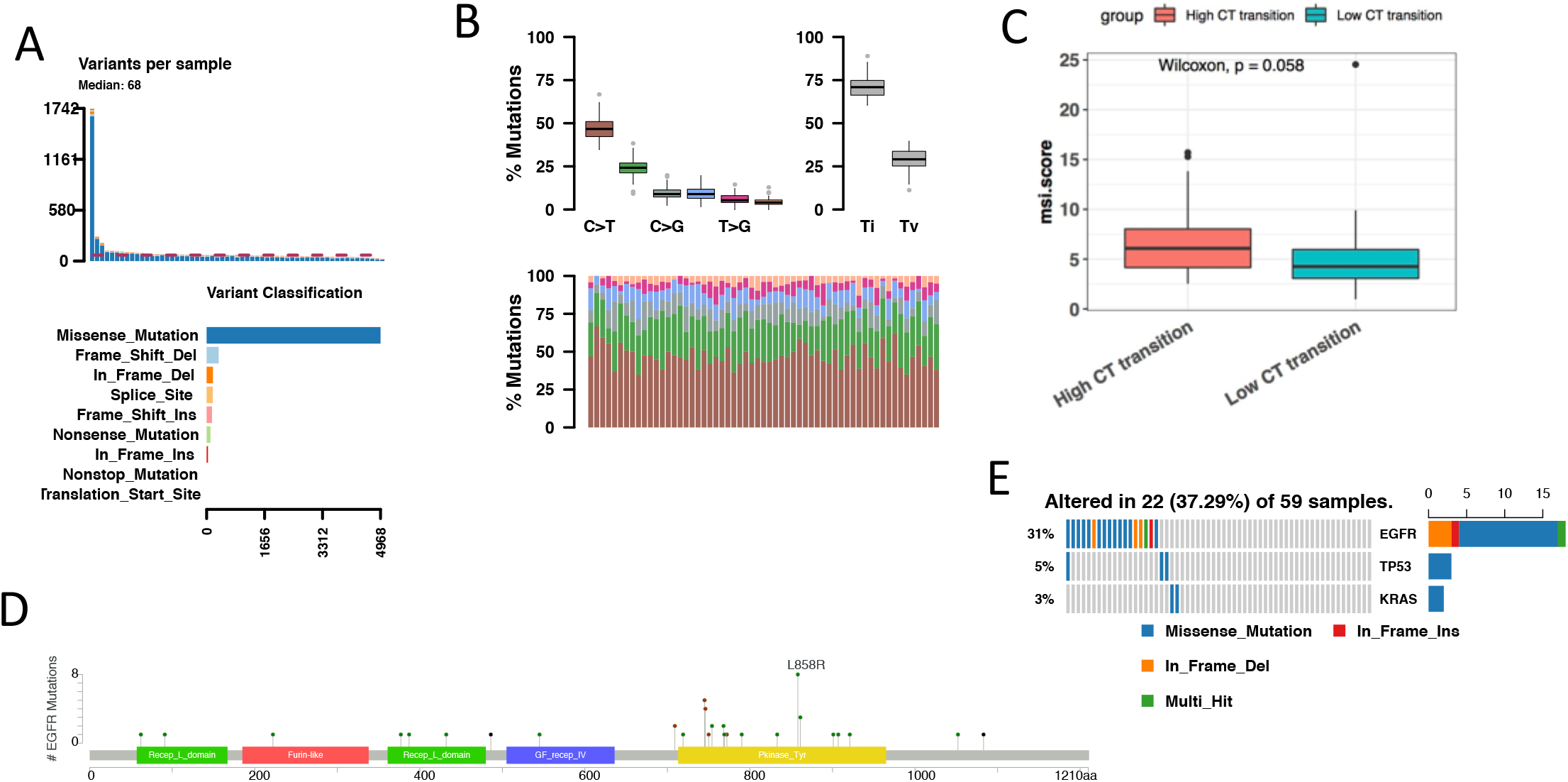
Mutation overview of MIA. The summary of variants from MIA, including the number of different variant classifications and SNV classes for each patient (**A**) and the frequency of nucleotide changes (**B**). (**C**) Comparison of msi score between high and low CT transition. (**D**) The mutation loci in EGFR. The mutation pattern of EGFR, KRAS and TP53 in MIA.

**Figure S5.**
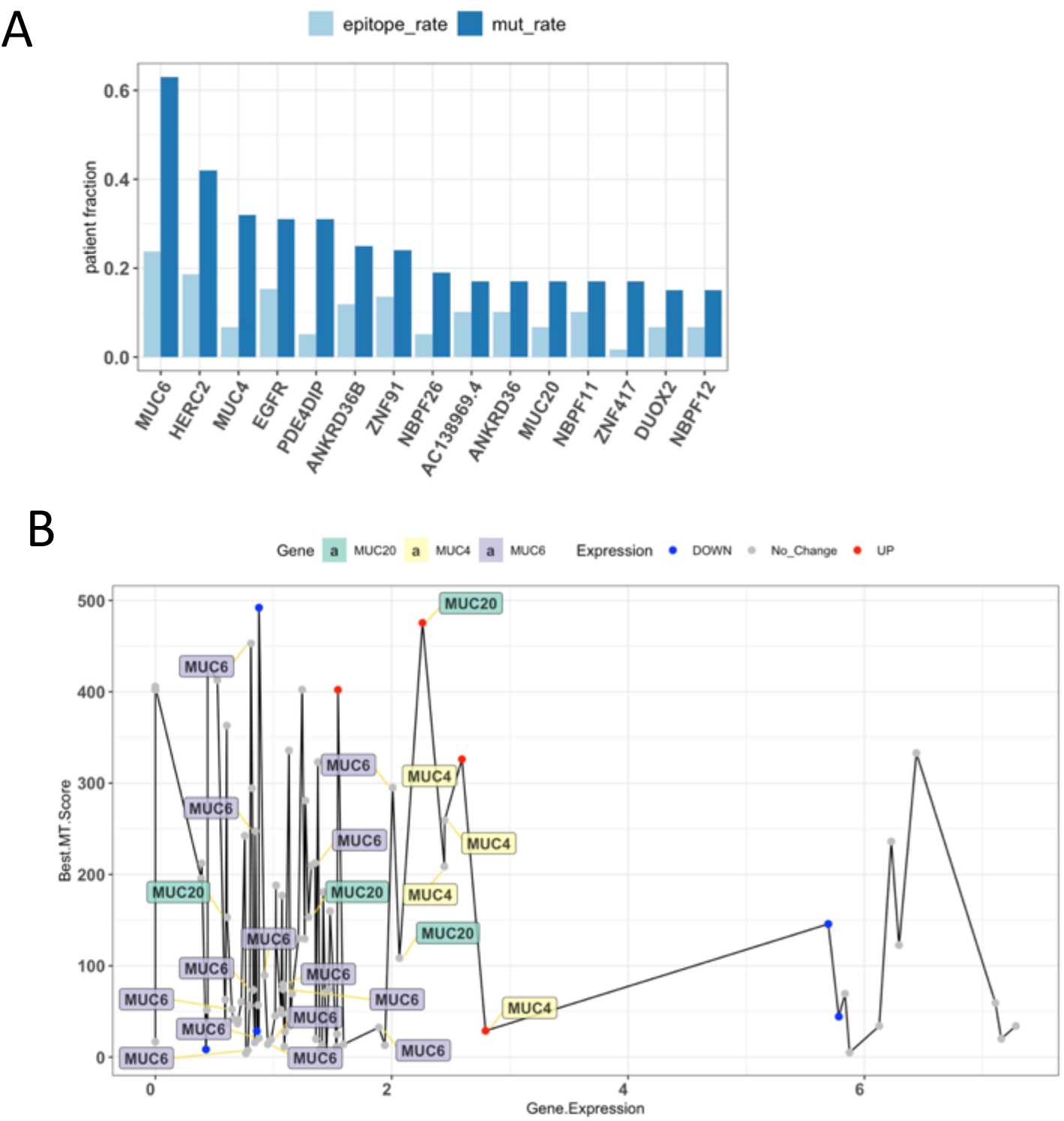
Neo-epitope prediction for top mutated genes. (**A**) Mutation rate (dark blue bar) and neo-epitope rate (MHC-I bound, light blue bar) among 59 patients for top 15 mutate genes. (**B**) Neo-epitope binding affinity and gene expression for top 15 mutated genes in patients. Each dot represents a predicted neo-epitope in a patient. Epitopes predicted from MUC20 (green), MUC4 (yellow), and MUC6 (purple) are highlighted with text. The color of dot represents the types of gene expression changes, including up-regulation (red), down-regulation (blue) and no change (grey).

**Figure S6.**
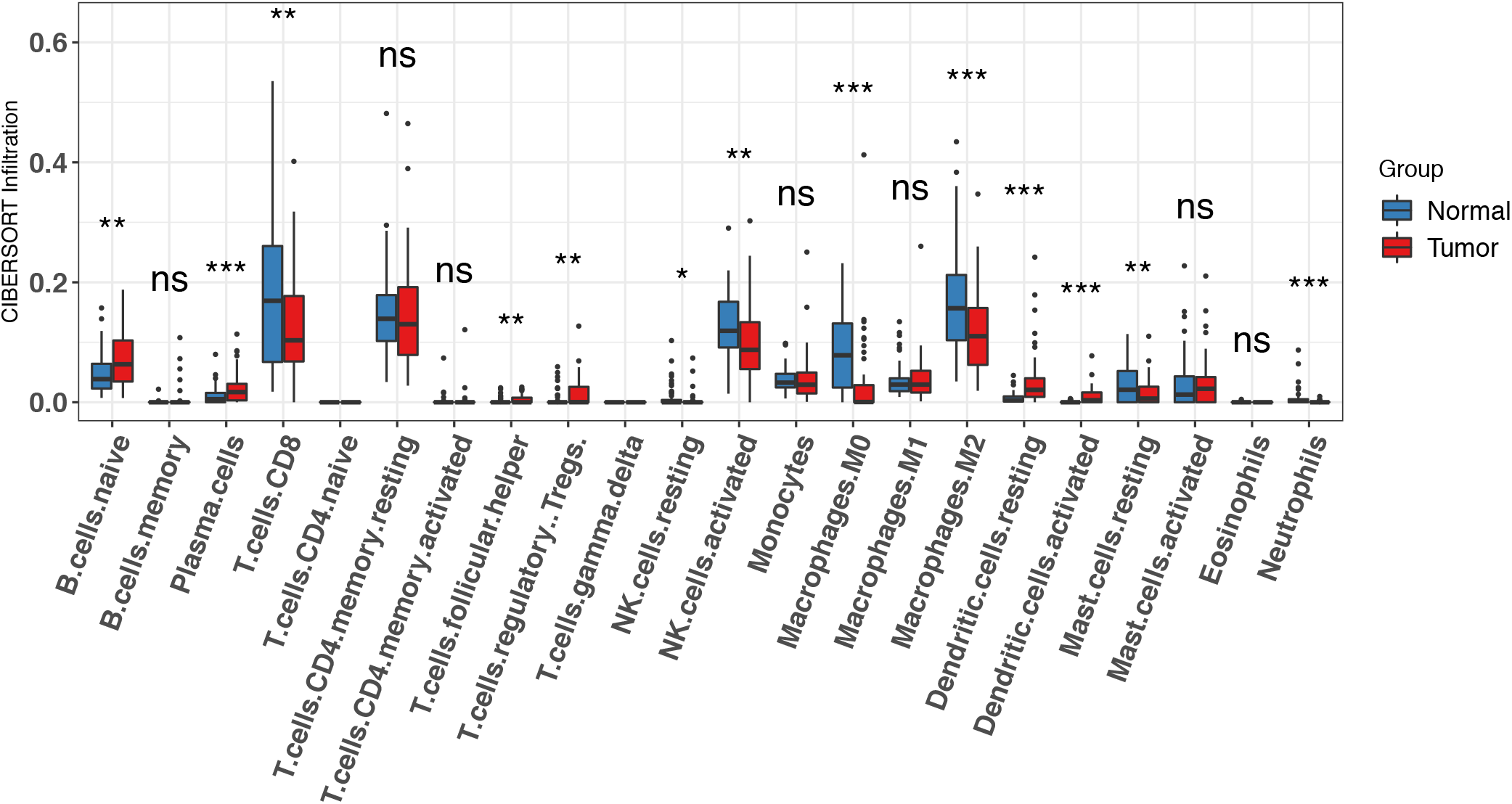
Immune infiltration estimation from CIBERSORT. Comparison of Immune cell infiltration estimated from CIBERSORT (“Absolute” mode) between MIA tumor and adjacent normal tissues.

**Figure S7.**
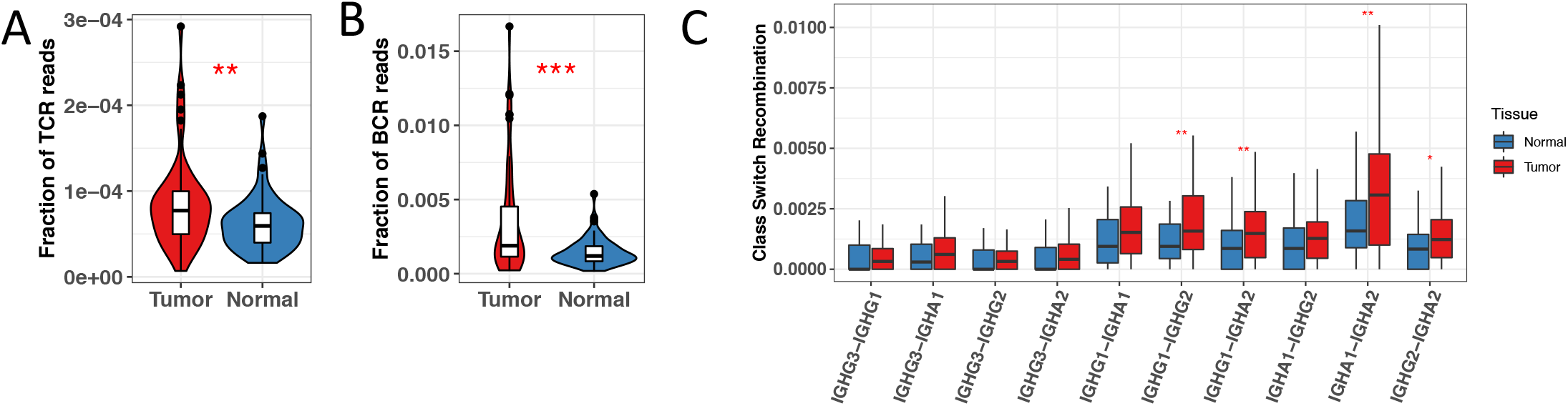
Immune repertoire metrics in MIA. Comparison of faction of TCR reads (**A**), faction of BCR reads (**B**) between MIA tumor and normal tissues. (**C**) The ratio of Ig class switch recombination between IgG and IgA in MIA tumor and normal tissues. (*** P < 0.001; ** P < 0.01; * P < 0.05).

**Figure S8.**
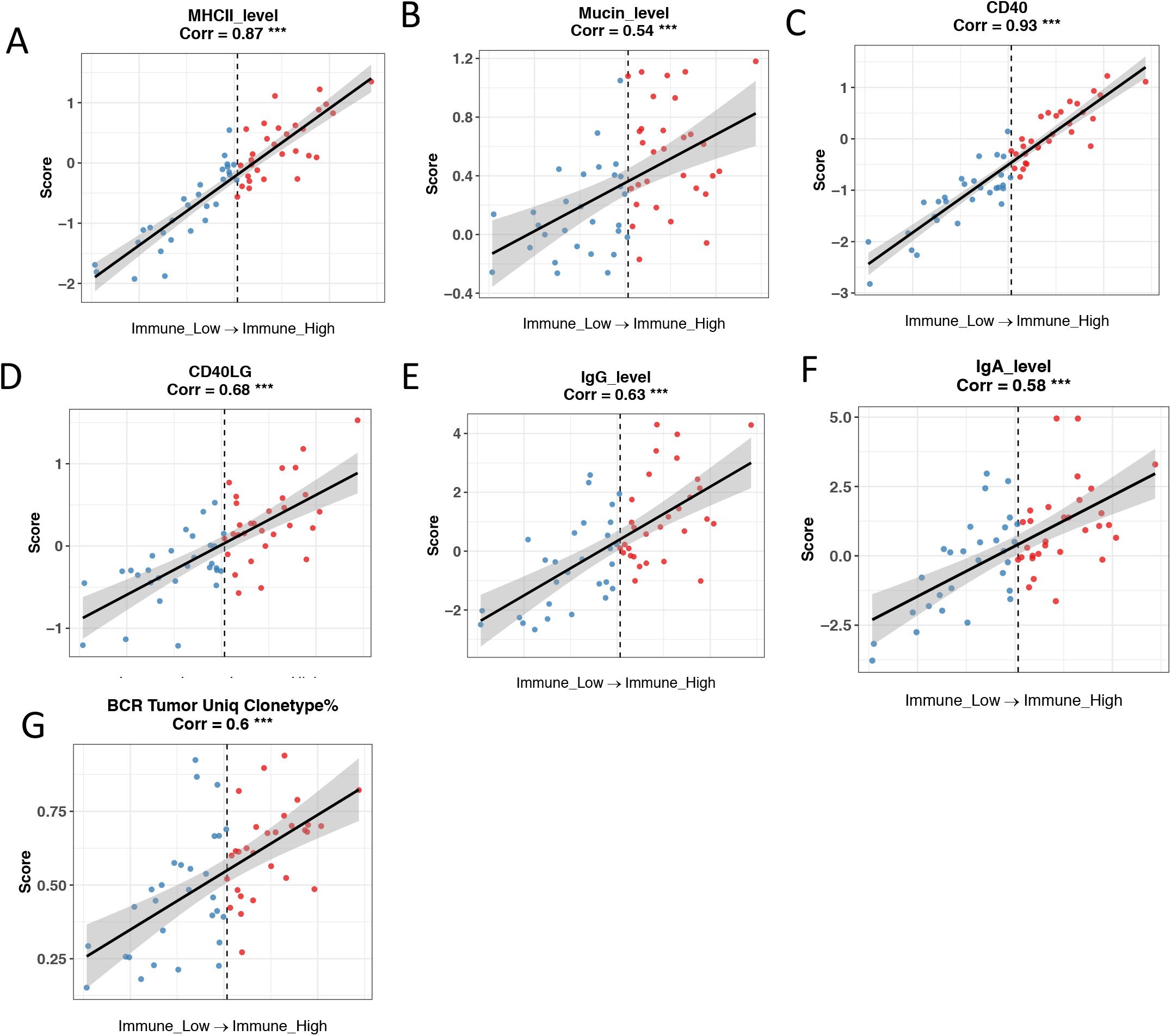
Association between molecules mediated B-CD4+ T activation and immune-related signature score. Association between MHCII level (**A**), mucin level (**B**), CD40 level (**C**), CD40LG level (**D**), IgG level (**E**), IgA level (**F**), and percentage of tumor unique BCR clonotypes (**G**) with immune-related signature score. MIA tumors are divided into “immune-low” and “immune-high” groups based on the median immune-related signature score. (*** P < 0.001; ** P < 0.01; * P < 0.05).

**Figure S9.**
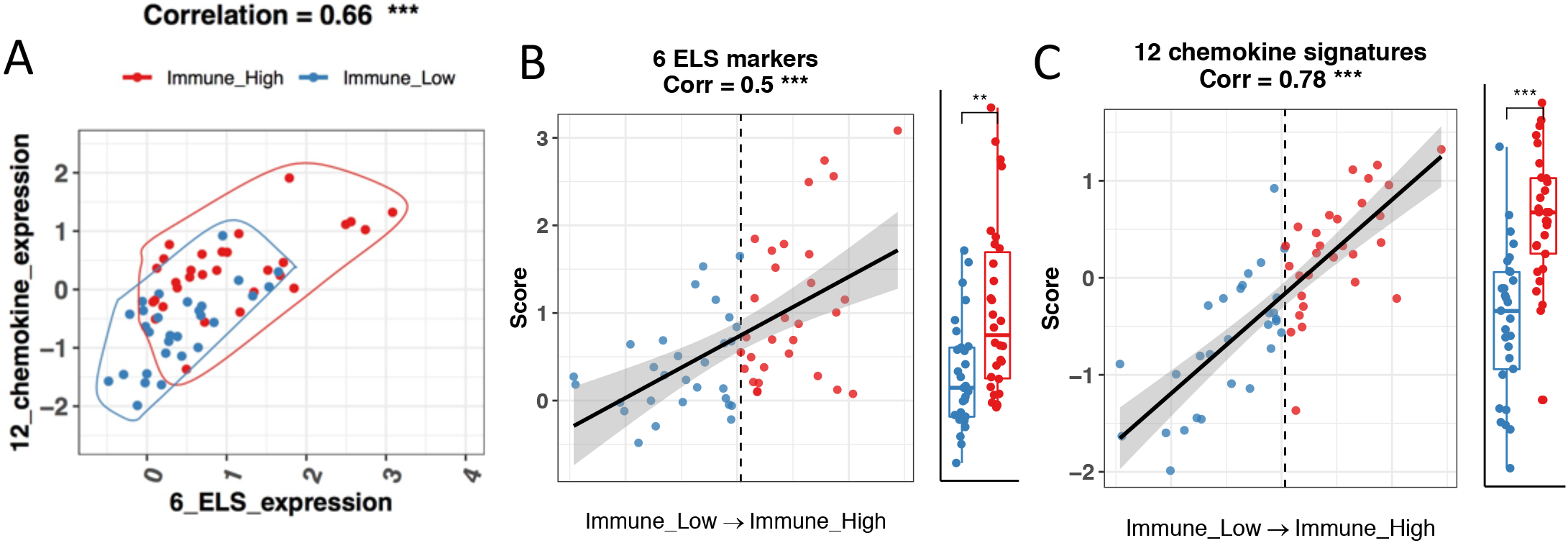
6 TLS markers and a 12-chemokine signature for predicting TLS existence. (**A**) Association between 6 TLS markers expression and a 12-chemokine signature score. (**B**) Association between 6 TLS markers expression and immune-related signature score. (**C**) Association between a 12-chemokine signature score and immune-related signature score. MIA tumors are divided into “immune-low” and “immune-high” groups based on the median immune-related signature score. (*** P < 0.001; ** P < 0.01; * P < 0.05).

**Figure S10.**
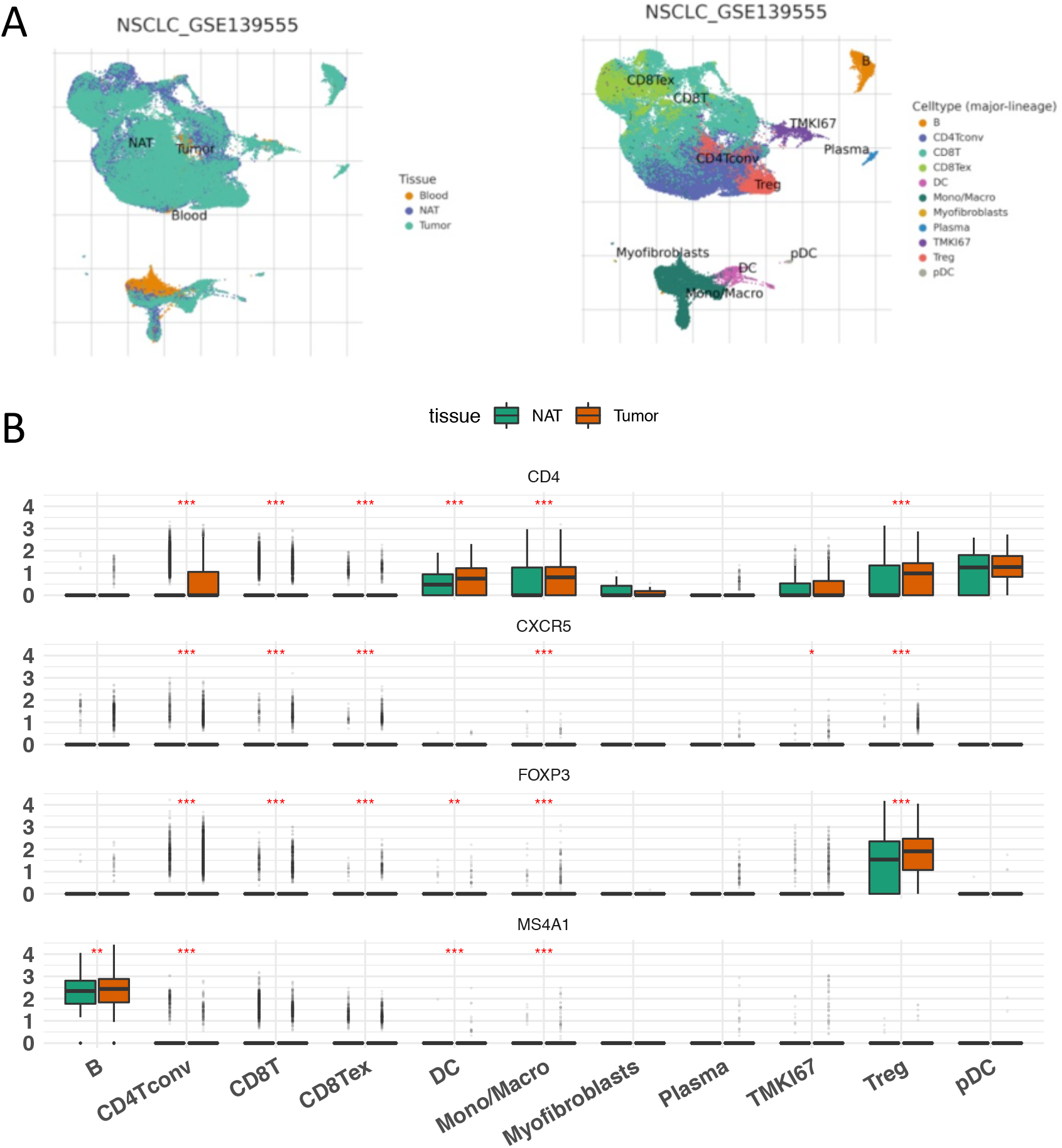
B, follicular T helper, and follicular T regulatory cells are enriched in NSCLC tumors in an independent single-cell RNA-seq dataset NSCLC_GSE139555. (**A**) (left) cell distribution in different tissues and each color represents a type of tissue (left), including blood, NAT (normal adjacent tissue), and tumor;(right) cell type annotation and each color represents a cell type. (**B**) Gene expressions for CD4, CXCR5, FOXP3, and MS4A1 are compared between tumor and NAT across different cell types. (*** P < 0.001; ** P < 0.01; * P < 0.05).

**Figure S11.**
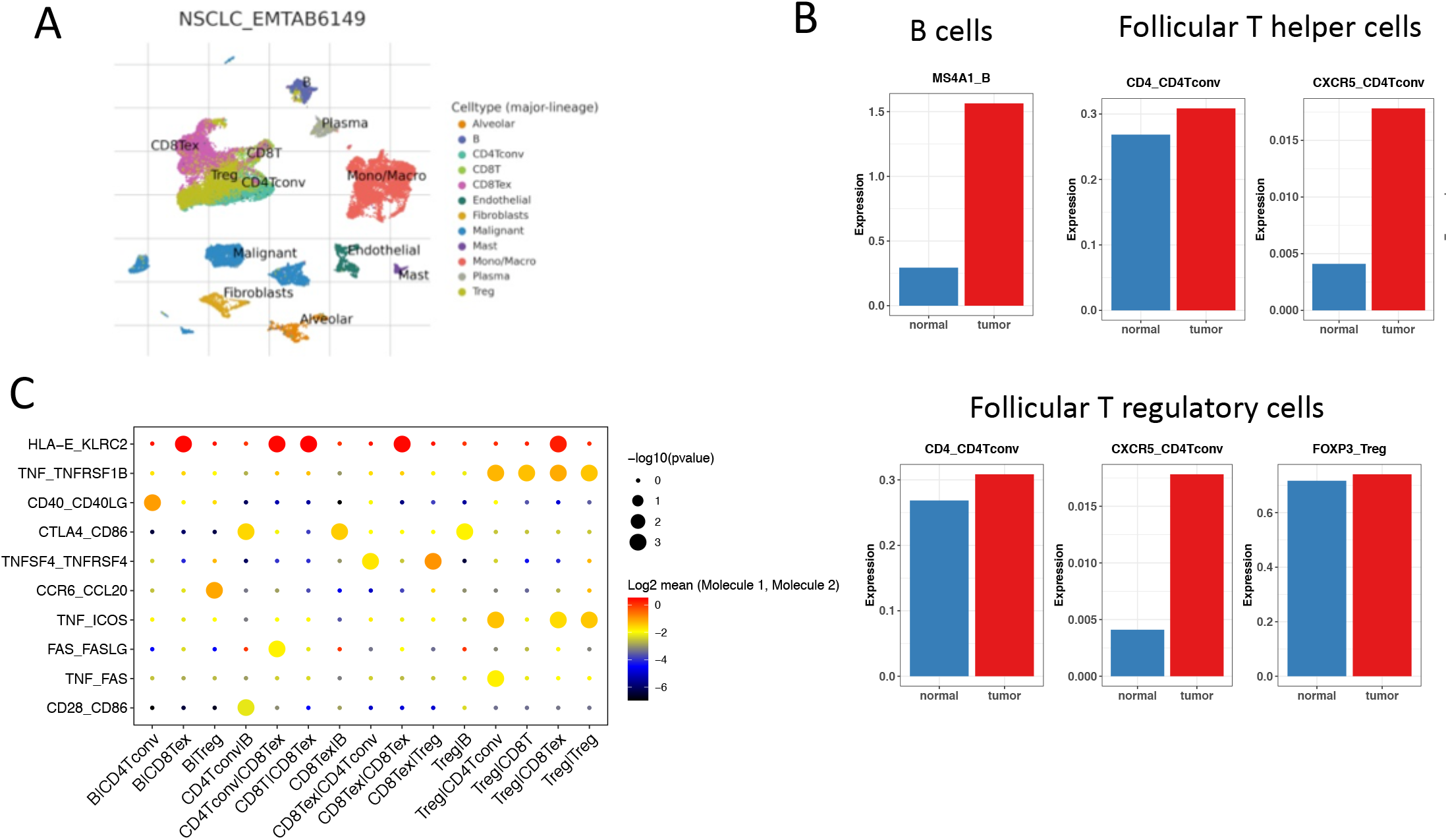
B, follicular T helper, and follicular T regulatory cells are enriched in NSCLC tumors in an independent single-cell RNA-seq dataset NSCLC_EMTAB6149. (**A**) cell type annotations. Each color represents a cell type. (**B**) Cluster averaged signature expression comparison between tumor (red) and normal tissues (blue) of different cell types. (**C**) Receptor-ligand pairs enrichment of different cell-cell interactions. The dot color represents the expression level of the receptor-ligand pairs and the dot size represents the enrichment significance of interaction pairs.

**Figure S12.**
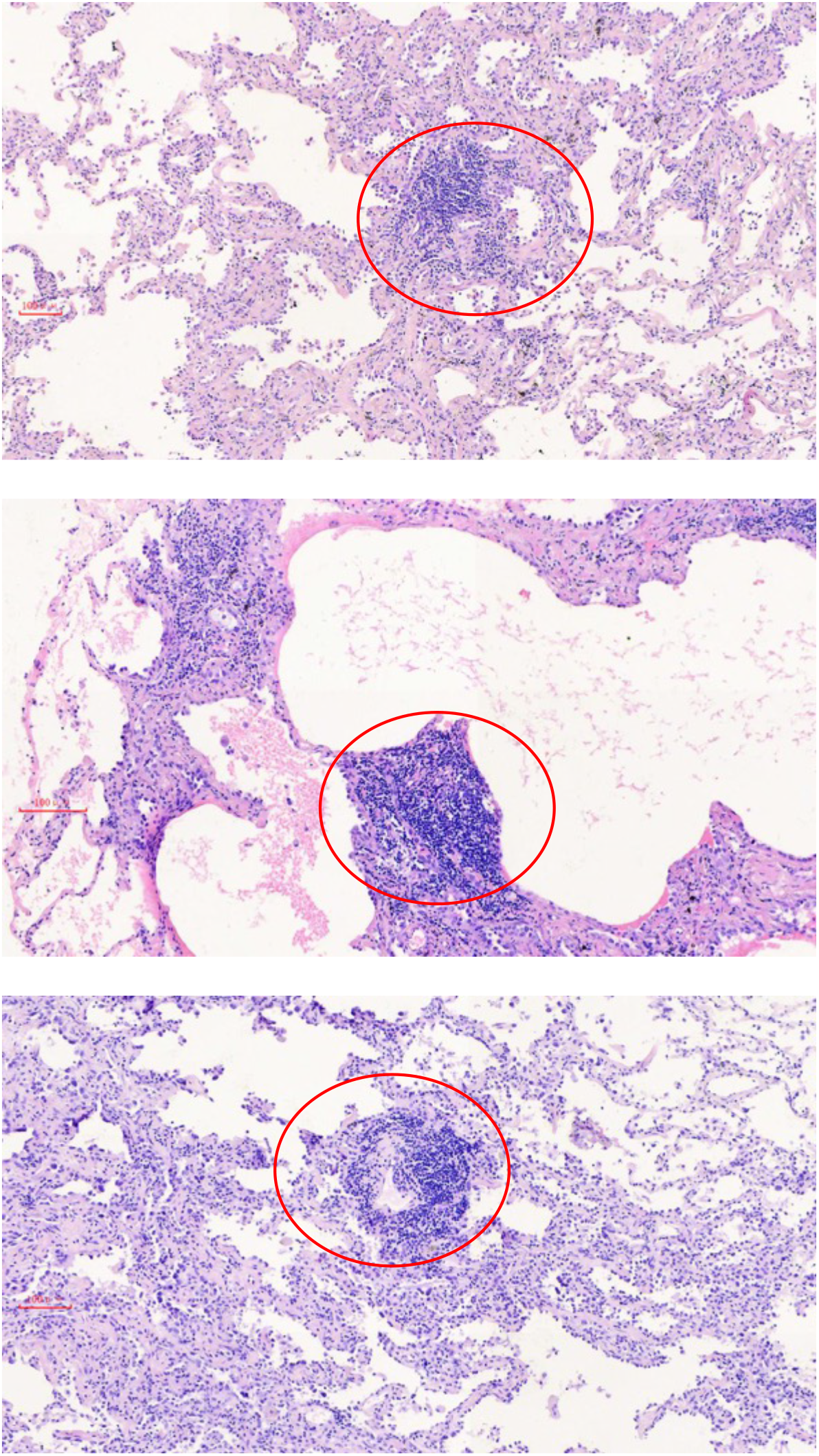
Evidence of TLS presence from H&E staining. The red circles denote the inferred TLS B and T cell aggregation.

**Figure S13.**
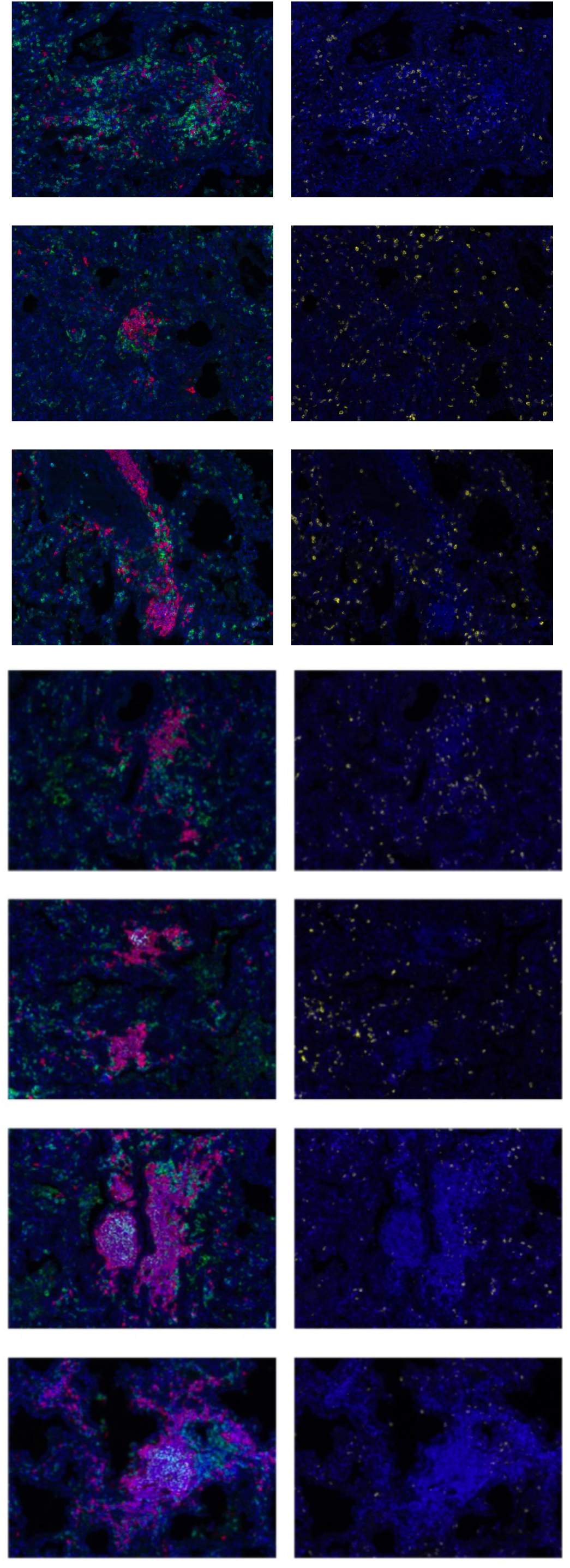
Different maturation level of TLS formation and corresponding CD8+ T cell infiltration. TLS formation stained by CD4(green), CD20(red), and CD35(cyan) co-localization in the left panel and CD8+ T cell infiltration in the right panel from the same observation field are shown.

**Figure S14.**
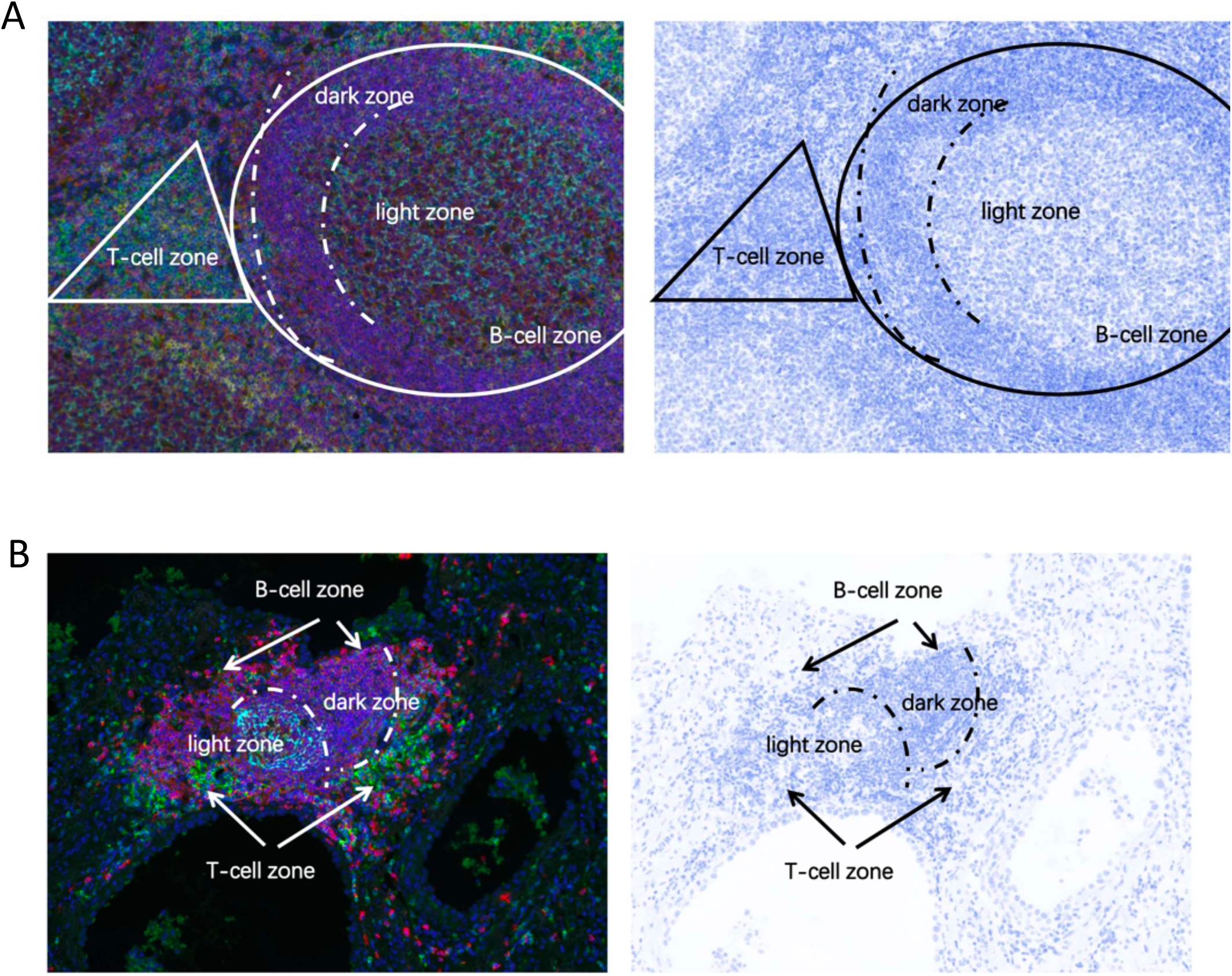
Normal germinal center in tonsil and ectopic germinal center in MIA. Normal germinal center in tonsil (**A**) and ectopic germinal center in MIA (**B**). T-cell zone, B-cell zone, dark zone, and light zone consist of GC in tonsil or EGC in MIA stained by immunofluorescence (left) and immunohistochemistry (right).

**Figure S15.**
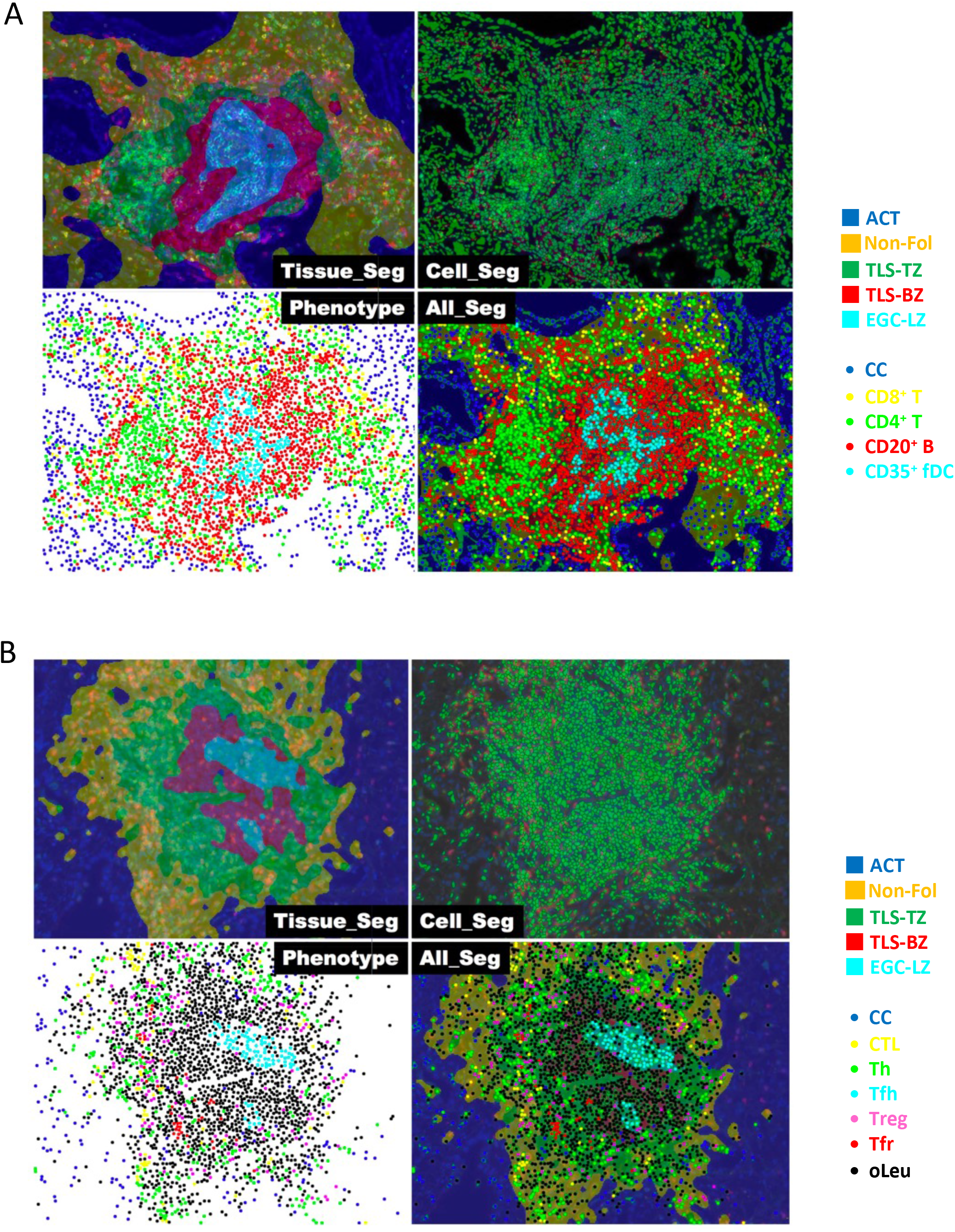
Multispectral analysis for segmenting tissues, cells and phenotypes. Segmentation for six-plex panel for TLS (**A**) and seven-plex panel for Tfr and Tfh (**B**). Each observation field could be segmented into 5 tissue categories as adenocarcinoma tissue (ACT), Non-Follicle area (Non-Fol), TLS T-cell area (TLS-T), TLS B-cell area (TLS-B) and light zone of germinal center (EGC-LZ) in both six-plex panel (top) and seven-plex panel (bottom). Five phenotypes were contained in six-plex panel, including CC (blue), CD8+ T (yellow), CD4+ T (green), CD20+ B (red), and CD35+ FDC (cyan). Seven phenotypes were contained in seven-plex, including CC (blue), CTL (yellow), Th (green), Tfh (cyan), Treg (pink), Tfr (red), and oLeu (black). ACC: Adenocarcinoma cancer cell; CC: cancer cell; Non-Fol: Non-follicular area; CTL: cytotoxic T lymphocyte; Th: T helper; Tfh: follicular T helper; Treg: regulatory T-cell; Tfr: follicular Treg; FDC: follicular dendritic cell; oLeu: other leukocyte.

**Figure S16.**
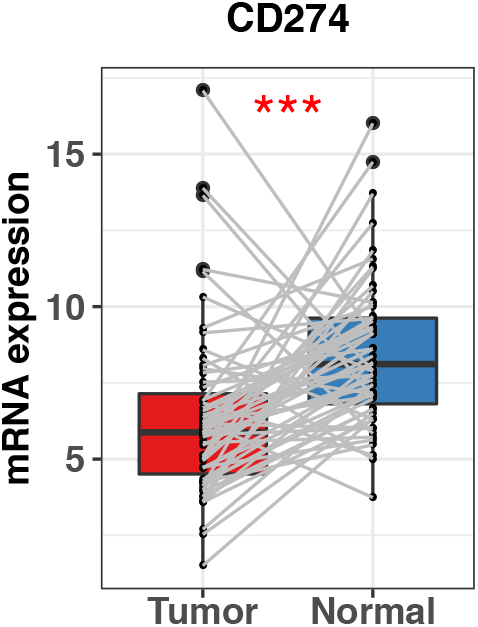
Down-regulated PD-L1 in MIA tumors. Comparison of the expression of PD-L1 (CD274) in tumor (red) and adjacent-normal (blue) samples (*** P < 0.001; ** P < 0.01; * P < 0.05).

**Figure S17.**
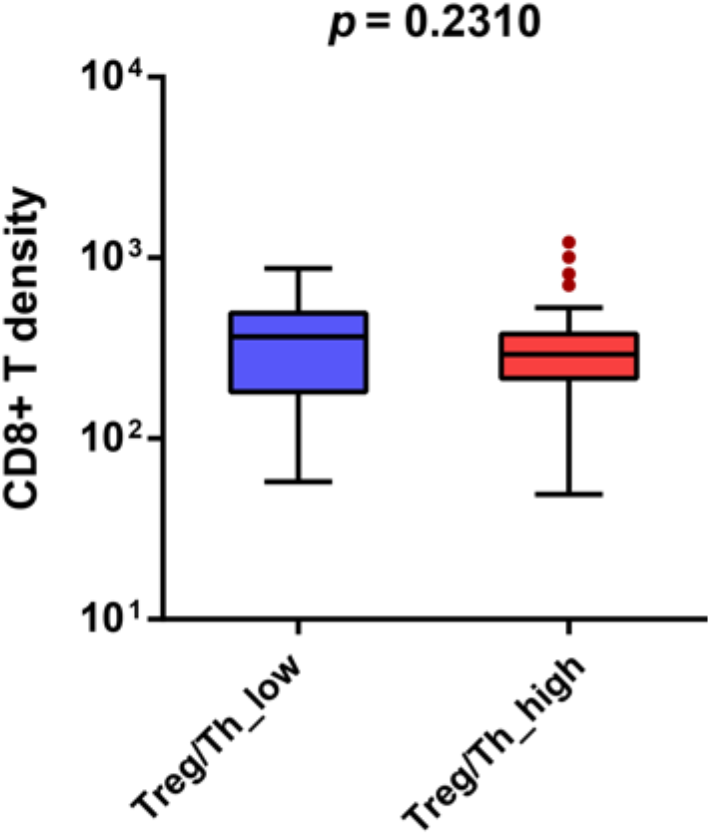
The ratio of regulatory T to helper T and CD8+ T cell infiltration in Non-follicle areas. CD8+ T cell comparison between groups with high (red) and low (blue) ratio of regulatory T to helper T.

